## Supplemental Appendices for "COVID-19 Increases the Rate of Incident Diabetes: A Case-Control Cohort Time-to-Event Study"

Shannon C. Phillips, MPH (1,2)

Kimberley D. Lucas, MPH (3)

Donna A. Jacobsen, DO (3)

David M. Studdert, LLB, ScD, MPH (1,2,4)

(1) Department of Health Policy, Stanford Medical School, Stanford University, Stanford, CA, 94306 USA

(2) Center for Health Policy, Freeman Spogli Institute, Stanford University, Stanford. CA, 94306 USA

(3) California Correctional Health Care Services, Elk Grove, CA, 95758 USA

(4) Stanford Law School, Stanford University, Stanford, CA, 94306 USA

**S1. Confounding Medications and Diagnoses**

Based on consultations with clinical subject matter experts, medications that can alter blood glucose were identified to enable the determination of clinical events (prescriptions of these medications) that could confound diagnoses of diabetes.

| **GCN Code** | **Description** |
| --- | --- |
| 6758 | betamethasone acetate/betamethasone sodium phosphate |
| 25750, 70506 | budesonide |
| 6780, 6781, 6782, 6784, 6785, 6786, 6787, 6788, 6789, 6790 | dexamethasone |
| 6776, 6778 | dexamethasone sodium phosphate |
| 62053 | dexamethasone sodium phosphate/PF |
| 37045, 6703, 6704, 6705 | hydrocortisone |
| 37045, 6703, 6704, 6705 | hydrocortisone |
| 45311, 6737, 6738, 6741, 6742 | methylprednisolone |
| 6724, 6725 | methylprednisolone acetate |
| 51554, 51555, 51557, 6729, 6730, 6732, 6733 | methylprednisolone sodium succinate |
| 65977, 65978, 65980 | methylprednisolone sodium succinate/PF |
| 45267, 45268, 6746, 6748, 6749, 6750, 6751, 6753, 6754 | prednisone |

Likewise, lists of diagnoses that would confound the analysis as they can lead to alterations in glucose levels were also identified via expert consultation.

| Code (SNOMed/ICD-10) | Description |
| --- | --- |
| 102660008 / E88.1 | Glucose level outside reference range |
| 237602007 | Metabolic Syndrome X (Insulin Resistance Syndrome) |
| 267433009 / E78.1 | Pure Hyperglyceridemia |
| 69878008 / E28.2 | Polycystic Ovary Syndrome |

**Appendix Figure 1. Flow Diagram of Inclusion in the Case-Control Cohort Time-to-Event Study**

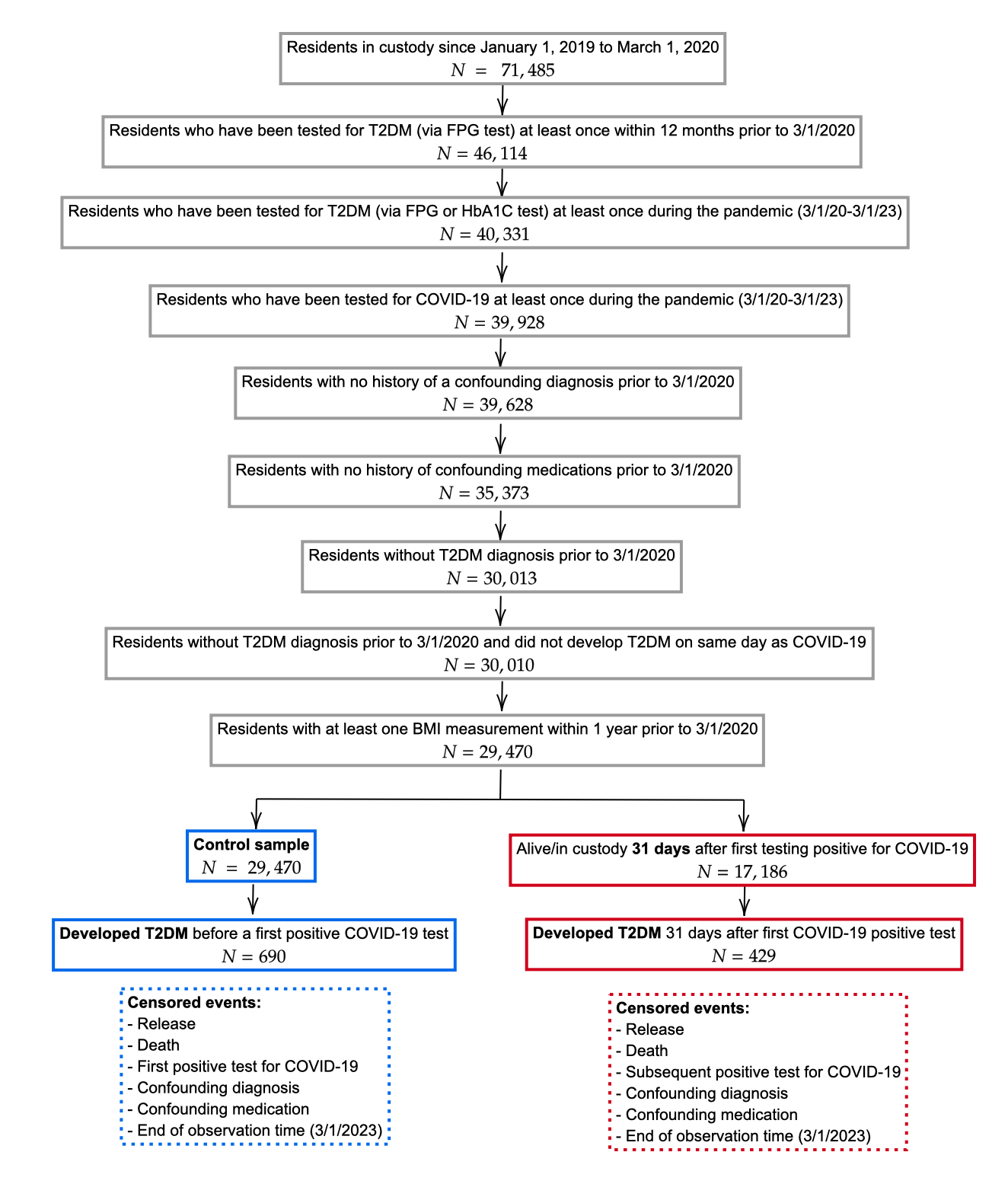

The figure shows the inclusion/exclusion criteria for individual in the study cohort. All cohort members began by contributing (non-exposure) observation time. Individuals who had their first positive COVID-19 test prior to being diagnosed with T2DM (a subset of the control/non-exposure sample) began contributing post-exposure time at 31 days after this test sample was collected (we excluded the first 30 days from the analysis).

**Appendix S2. Diabetes Diagnostic Codes**

Individuals were considered to have a diabetes diagnosis if their electronic health record contained any of the following codes. Their diagnosis date was determined as the earliest date at which one of the codes appeared in their health record.

| **Code Type** | **Code** | **Description** |
| --- | --- | --- |
| ICD-10 | E08 | Diabetes mellitus due to underlying condition |
| ICD-10 | E08.00 | Diabetes mellitus due to underlying condition with hyperosmolarity without nonketotic hyperglycemic-hyperosmolar coma (NKHHC) |
| ICD-10 | E08.2 | Diabetes mellitus due to underlying condition with kidney complications |
| ICD-10 | E08.21 | Diabetes mellitus due to underlying condition with diabetic nephropathy |
| ICD-10 | E08.22 | Diabetes mellitus due to underlying condition with diabetic chronic kidney disease |
| ICD-10 | E08.29 | Diabetes mellitus due to underlying condition with other diabetic kidney complication |
| ICD-10 | E08.3 | Diabetes mellitus due to underlying condition with ophthalmic complications |
| ICD-10 | E08.31 | Diabetes mellitus due to underlying condition with unspecified diabetic retinopathy |
| ICD-10 | E08.311 | Diabetes mellitus due to underlying condition with unspecified diabetic retinopathy with macular edema |
| ICD-10 | E08.319 | Diabetes mellitus due to underlying condition with unspecified diabetic retinopathy without macular edema |
| ICD-10 | E08.32 | Diabetes mellitus due to underlying condition with mild nonproliferative diabetic retinopathy |
| ICD-10 | E08.321 | Diabetes mellitus due to underlying condition with mild nonproliferative diabetic retinopathy with macular edema |
| ICD-10 | E08.3212 | Diabetes mellitus due to underlying condition with mild nonproliferative diabetic retinopathy with macular edema, left eye |
| ICD-10 | E08.329 | Diabetes mellitus due to underlying condition with mild nonproliferative diabetic retinopathy without macular edema |
| ICD-10 | E08.3291 | Diabetes mellitus due to underlying condition with mild nonproliferative diabetic retinopathy without macular edema, right eye |
| ICD-10 | E08.3292 | Diabetes mellitus due to underlying condition with mild nonproliferative diabetic retinopathy without macular edema, left eye |
| ICD-10 | E08.3293 | Diabetes mellitus due to underlying condition with mild nonproliferative diabetic retinopathy without macular edema, bilateral |
| ICD-10 | E08.3299 | Diabetes mellitus due to underlying condition with mild nonproliferative diabetic retinopathy without macular edema, unspecified eye |
| ICD-10 | E08.33 | Diabetes mellitus due to underlying condition with moderate nonproliferative diabetic retinopathy |
| ICD-10 | E08.339 | Diabetes mellitus due to underlying condition with moderate nonproliferative diabetic retinopathy without macular edema |
| ICD-10 | E08.34 | Diabetes mellitus due to underlying condition with severe nonproliferative diabetic retinopathy |
| ICD-10 | E08.341 | Diabetes mellitus due to underlying condition with severe nonproliferative diabetic retinopathy with macular edema |
| ICD-10 | E08.35 | Diabetes mellitus due to underlying condition with proliferative diabetic retinopathy |
| ICD-10 | E08.351 | Diabetes mellitus due to underlying condition with proliferative diabetic retinopathy with macular edema |
| ICD-10 | E08.3513 | Diabetes mellitus due to underlying condition with proliferative diabetic retinopathy with macular edema, bilateral |
| ICD-10 | E08.3519 | Diabetes mellitus due to underlying condition with proliferative diabetic retinopathy with macular edema, unspecified eye |
| ICD-10 | E08.3553 | Diabetes mellitus due to underlying condition with stable proliferative diabetic retinopathy, bilateral |
| ICD-10 | E08.3592 | Diabetes mellitus due to underlying condition with proliferative diabetic retinopathy without macular edema, left eye |
| ICD-10 | E08.3593 | Diabetes mellitus due to underlying condition with proliferative diabetic retinopathy without macular edema, bilateral |
| ICD-10 | E08.37X2 | Diabetes mellitus due to underlying condition with diabetic macular edema, resolved following treatment, left eye |
| ICD-10 | E08.39 | Diabetes mellitus due to underlying condition with other diabetic ophthalmic complication |
| ICD-10 | E08.4 | Diabetes mellitus due to underlying condition with neurological complications |
| ICD-10 | E08.40 | Diabetes mellitus due to underlying condition with diabetic neuropathy, unspecified |
| ICD-10 | E08.41 | Diabetes mellitus due to underlying condition with diabetic mononeuropathy |
| ICD-10 | E08.42 | Diabetes mellitus due to underlying condition with diabetic polyneuropathy |
| ICD-10 | E08.43 | Diabetes mellitus due to underlying condition with diabetic autonomic (poly)neuropathy |
| ICD-10 | E08.49 | Diabetes mellitus due to underlying condition with other diabetic neurological complication |
| ICD-10 | E08.5 | Diabetes mellitus due to underlying condition with circulatory complications |
| ICD-10 | E08.59 | Diabetes mellitus due to underlying condition with other circulatory complications |
| ICD-10 | E08.6 | Diabetes mellitus due to underlying condition with other specified complications |
| ICD-10 | E08.61 | Diabetes mellitus due to underlying condition with diabetic arthropathy |
| ICD-10 | E08.610 | Diabetes mellitus due to underlying condition with diabetic neuropathic arthropathy |
| ICD-10 | E08.621 | Diabetes mellitus due to underlying condition with foot ulcer |
| ICD-10 | E08.622 | Diabetes mellitus due to underlying condition with other skin ulcer |
| ICD-10 | E08.628 | Diabetes mellitus due to underlying condition with other skin complications |
| ICD-10 | E08.649 | Diabetes mellitus due to underlying condition with hypoglycemia without coma |
| ICD-10 | E08.65 | Diabetes mellitus due to underlying condition with hyperglycemia |
| ICD-10 | E08.69 | Diabetes mellitus due to underlying condition with other specified complication |
| ICD-10 | E08.8 | Diabetes mellitus due to underlying condition with unspecified complications |
| ICD-10 | E08.9 | Diabetes mellitus due to underlying condition without complications |
| ICD-10 | E09.22 | Drug or chemical induced diabetes mellitus with diabetic chronic kidney disease |
| ICD-10 | E09.31 | Drug or chemical induced diabetes mellitus with unspecified diabetic retinopathy |
| ICD-10 | E09.319 | Drug or chemical induced diabetes mellitus with unspecified diabetic retinopathy without macular edema |
| ICD-10 | E09.359 | Drug or chemical induced diabetes mellitus with proliferative diabetic retinopathy without macular edema |
| ICD-10 | E09.9 | Drug or chemical induced diabetes mellitus without complications |
| ICD-10 | E10 | Type 1 diabetes mellitus |
| ICD-10 | E10.1 | Type 1 diabetes mellitus with acidosis |
| ICD-10 | E10.10 | Type 1 diabetes mellitus with ketoacidosis |
| ICD-10 | E10.10 | Type 1 diabetes mellitus with ketoacidosis without coma |
| ICD-10 | E10.21 | Type 1 diabetes mellitus with diabetic nephropathy |
| ICD-10 | E10.21 | Type 1 diabetes mellitus with established diabetic nephropathy |
| ICD-10 | E10.22 | Type 1 diabetes mellitus with end-stage renal disease [ESRD] |
| ICD-10 | E10.22 | Type 1 diabetes mellitus with diabetic chronic kidney disease |
| ICD-10 | E10.31 | Type 1 diabetes mellitus with preproliferative retinopathy |
| ICD-10 | E10.319 | Type 1 diabetes mellitus with unspecified diabetic retinopathy without macular edema |
| ICD-10 | E10.321 | Type 1 diabetes mellitus with mild nonproliferative diabetic retinopathy with macular edema |
| ICD-10 | E10.3213 | Type 1 diabetes mellitus with mild nonproliferative diabetic retinopathy with macular edema, bilateral |
| ICD-10 | E10.3219 | Type 1 diabetes mellitus with mild nonproliferative diabetic retinopathy with macular edema, unspecified eye |
| ICD-10 | E10.3293 | Type 1 diabetes mellitus with mild nonproliferative diabetic retinopathy without macular edema, bilateral |
| ICD-10 | E10.3299 | Type 1 diabetes mellitus with mild nonproliferative diabetic retinopathy without macular edema, unspecified eye |
| ICD-10 | E10.331 | Type 1 diabetes mellitus with moderate nonproliferative diabetic retinopathy with macular edema |
| ICD-10 | E10.339 | Type 1 diabetes mellitus with moderate nonproliferative diabetic retinopathy without macular edema |
| ICD-10 | E10.349 | Type 1 diabetes mellitus with severe nonproliferative diabetic retinopathy without macular edema |
| ICD-10 | E10.3519 | Type 1 diabetes mellitus with proliferative diabetic retinopathy with macular edema, unspecified eye |
| ICD-10 | E10.3599 | Type 1 diabetes mellitus with proliferative diabetic retinopathy without macular edema, unspecified eye |
| ICD-10 | E10.36 | Type 1 diabetes mellitus with diabetic cataract |
| ICD-10 | E10.37X9 | Type 1 diabetes mellitus with diabetic macular edema, resolved following treatment, unspecified eye |
| ICD-10 | E10.39 | Type 1 diabetes mellitus with other diabetic ophthalmic complication |
| ICD-10 | E10.4 | Type 1 diabetes mellitus with neurological complications |
| ICD-10 | E10.40 | Type 1 diabetes mellitus with diabetic neuropathy, unspecified |
| ICD-10 | E10.40 | Type 1 diabetes mellitus with mononeuropathy |
| ICD-10 | E10.41 | Type 1 diabetes mellitus with polyneuropathy |
| ICD-10 | E10.42 | Type 1 diabetes mellitus with autonomic neuropathy |
| ICD-10 | E10.42 | Type 1 diabetes mellitus with diabetic polyneuropathy |
| ICD-10 | E10.43 | Type 1 diabetes mellitus with diabetic autonomic (poly)neuropathy |
| ICD-10 | E10.44 | Type 1 diabetes mellitus with diabetic amyotrophy |
| ICD-10 | E10.49 | Type 1 diabetes mellitus with other diabetic neurological complication |
| ICD-10 | E10.5 | Type 1 diabetes mellitus with circulatory complications |
| ICD-10 | E10.59 | Type 1 diabetes mellitus with other circulatory complications |
| ICD-10 | E10.610 | Type 1 diabetes mellitus with diabetic neuropathic arthropathy |
| ICD-10 | E10.621 | Type 1 diabetes mellitus with foot ulcer |
| ICD-10 | E10.622 | Type 1 diabetes mellitus with other skin ulcer |
| ICD-10 | E10.64 | Type 1 diabetes mellitus with poor control, so described |
| ICD-10 | E10.649 | Type 1 diabetes mellitus with hypoglycemia without coma |
| ICD-10 | E10.65 | Type 1 diabetes mellitus with hyperglycemia |
| ICD-10 | E10.69 | Type 1 diabetes mellitus with other specified complication |
| ICD-10 | E10.8 | Type 1 diabetes mellitus with unspecified complications |
| ICD-10 | E10.9 | Type 1 diabetes mellitus without complications |
| ICD-10 | E10.9 | Type 1 diabetes mellitus without (mention of) complication |
| ICD-10 | E11 | Type 2 diabetes mellitus |
| ICD-10 | E11.00 | Type 2 diabetes mellitus with hyperosmolarity without nonketotic hyperglycemic-hyperosmolar coma (NKHHC) |
| ICD-10 | E11.01 | Type 2 diabetes mellitus with hyperosmolarity with coma |
| ICD-10 | E11.10 | Type 2 diabetes mellitus with ketoacidosis without coma |
| ICD-10 | E11.10 | Type 2 diabetes mellitus with ketoacidosis |
| ICD-10 | E11.21 | Type 2 diabetes mellitus with diabetic nephropathy |
| ICD-10 | E11.21 | Type 2 diabetes mellitus with established diabetic nephropathy |
| ICD-10 | E11.22 | Type 2 diabetes mellitus with diabetic chronic kidney disease |
| ICD-10 | E11.22 | Type 2 diabetes mellitus with end-stage renal disease [ESRD] |
| ICD-10 | E11.29 | Type 2 diabetes mellitus with other diabetic kidney complication |
| ICD-10 | E11.31 | Type 2 diabetes mellitus with preproliferative retinopathy |
| ICD-10 | E11.311 | Type 2 diabetes mellitus with unspecified diabetic retinopathy with macular edema |
| ICD-10 | E11.319 | Type 2 diabetes mellitus with unspecified diabetic retinopathy without macular edema |
| ICD-10 | E11.32 | Type 2 diabetes mellitus with proliferative retinopathy |
| ICD-10 | E11.321 | Type 2 diabetes mellitus with mild nonproliferative diabetic retinopathy with macular edema |
| ICD-10 | E11.3211 | Type 2 diabetes mellitus with mild nonproliferative diabetic retinopathy with macular edema, right eye |
| ICD-10 | E11.3213 | Type 2 diabetes mellitus with mild nonproliferative diabetic retinopathy with macular edema, bilateral |
| ICD-10 | E11.3219 | Type 2 diabetes mellitus with mild nonproliferative diabetic retinopathy with macular edema, unspecified eye |
| ICD-10 | E11.329 | Type 2 diabetes mellitus with mild nonproliferative diabetic retinopathy without macular edema |
| ICD-10 | E11.3291 | Type 2 diabetes mellitus with mild nonproliferative diabetic retinopathy without macular edema, right eye |
| ICD-10 | E11.3292 | Type 2 diabetes mellitus with mild nonproliferative diabetic retinopathy without macular edema, left eye |
| ICD-10 | E11.3293 | Type 2 diabetes mellitus with mild nonproliferative diabetic retinopathy without macular edema, bilateral |
| ICD-10 | E11.3299 | Type 2 diabetes mellitus with mild nonproliferative diabetic retinopathy without macular edema, unspecified eye |
| ICD-10 | E11.331 | Type 2 diabetes mellitus with moderate nonproliferative diabetic retinopathy with macular edema |
| ICD-10 | E11.339 | Type 2 diabetes mellitus with moderate nonproliferative diabetic retinopathy without macular edema |
| ICD-10 | E11.3393 | Type 2 diabetes mellitus with moderate nonproliferative diabetic retinopathy without macular edema, bilateral |
| ICD-10 | E11.3399 | Type 2 diabetes mellitus with moderate nonproliferative diabetic retinopathy without macular edema, unspecified eye |
| ICD-10 | E11.34 | Type 2 diabetes mellitus with severe nonproliferative diabetic retinopathy |
| ICD-10 | E11.3419 | Type 2 diabetes mellitus with severe nonproliferative diabetic retinopathy with macular edema, unspecified eye |
| ICD-10 | E11.3499 | Type 2 diabetes mellitus with severe nonproliferative diabetic retinopathy without macular edema, unspecified eye |
| ICD-10 | E11.351 | Type 2 diabetes mellitus with proliferative diabetic retinopathy with macular edema |
| ICD-10 | E11.3511 | Type 2 diabetes mellitus with proliferative diabetic retinopathy with macular edema, right eye |
| ICD-10 | E11.3519 | Type 2 diabetes mellitus with proliferative diabetic retinopathy with macular edema, unspecified eye |
| ICD-10 | E11.3521 | Type 2 diabetes mellitus with proliferative diabetic retinopathy with traction retinal detachment involving the macula, right eye |
| ICD-10 | E11.3523 | Type 2 diabetes mellitus with proliferative diabetic retinopathy with traction retinal detachment involving the macula, bilateral |
| ICD-10 | E11.3539 | Type 2 diabetes mellitus with proliferative diabetic retinopathy with traction retinal detachment not involving the macula, unspecified eye |
| ICD-10 | E11.3553 | Type 2 diabetes mellitus with stable proliferative diabetic retinopathy, bilateral |
| ICD-10 | E11.359 | Type 2 diabetes mellitus with proliferative diabetic retinopathy without macular edema |
| ICD-10 | E11.3592 | Type 2 diabetes mellitus with proliferative diabetic retinopathy without macular edema, left eye |
| ICD-10 | E11.3593 | Type 2 diabetes mellitus with proliferative diabetic retinopathy without macular edema, bilateral |
| ICD-10 | E11.3599 | Type 2 diabetes mellitus with proliferative diabetic retinopathy without macular edema, unspecified eye |
| ICD-10 | E11.36 | Type 2 diabetes mellitus with diabetic cataract |
| ICD-10 | E11.36 | Type 2 diabetes mellitus with advanced ophthalmic disease |
| ICD-10 | E11.37X9 | Type 2 diabetes mellitus with diabetic macular edema, resolved following treatment, unspecified eye |
| ICD-10 | E11.39 | Type 2 diabetes mellitus with other diabetic ophthalmic complication |
| ICD-10 | E11.40 | Type 2 diabetes mellitus with mononeuropathy |
| ICD-10 | E11.40 | Type 2 diabetes mellitus with diabetic neuropathy, unspecified |
| ICD-10 | E11.41 | Type 2 diabetes mellitus with diabetic mononeuropathy |
| ICD-10 | E11.41 | Type 2 diabetes mellitus with polyneuropathy |
| ICD-10 | E11.42 | Type 2 diabetes mellitus with autonomic neuropathy |
| ICD-10 | E11.42 | Type 2 diabetes mellitus with diabetic polyneuropathy |
| ICD-10 | E11.43 | Type 2 diabetes mellitus with diabetic autonomic (poly)neuropathy |
| ICD-10 | E11.44 | Type 2 diabetes mellitus with diabetic amyotrophy |
| ICD-10 | E11.49 | Type 2 diabetes mellitus with other diabetic neurological complication |
| ICD-10 | E11.5 | Type 2 diabetes mellitus with circulatory complications |
| ICD-10 | E11.51 | Type 2 diabetes mellitus with diabetic peripheral angiopathy without gangrene |
| ICD-10 | E11.51 | Type 2 diabetes mellitus with peripheral angiopathy with gangrene |
| ICD-10 | E11.52 | Type 2 diabetes mellitus with certain circulatory complications |
| ICD-10 | E11.59 | Type 2 diabetes mellitus with other circulatory complications |
| ICD-10 | E11.6 | Type 2 diabetes mellitus with other specified complications |
| ICD-10 | E11.610 | Type 2 diabetes mellitus with diabetic neuropathic arthropathy |
| ICD-10 | E11.618 | Type 2 diabetes mellitus with other diabetic arthropathy |
| ICD-10 | E11.62 | Type 2 diabetes mellitus with periodontal complication |
| ICD-10 | E11.620 | Type 2 diabetes mellitus with diabetic dermatitis |
| ICD-10 | E11.621 | Type 2 diabetes mellitus with foot ulcer |
| ICD-10 | E11.622 | Type 2 diabetes mellitus with other skin ulcer |
| ICD-10 | E11.628 | Type 2 diabetes mellitus with other skin complications |
| ICD-10 | E11.638 | Type 2 diabetes mellitus with other oral complications |
| ICD-10 | E11.64 | Type 2 diabetes mellitus with poor control, so described |
| ICD-10 | E11.641 | Type 2 diabetes mellitus with hypoglycemia with coma |
| ICD-10 | E11.649 | Type 2 diabetes mellitus with hypoglycemia without coma |
| ICD-10 | E11.65 | Type 2 diabetes mellitus with hyperglycemia |
| ICD-10 | E11.69 | Type 2 diabetes mellitus with other specified complication |
| ICD-10 | E11.8 | Type 2 diabetes mellitus with unspecified complications |
| ICD-10 | E11.9 | Type 2 diabetes mellitus without (mention of) complications |
| ICD-10 | E11.9 | Type 2 diabetes mellitus without complications |
| ICD-10 | E13 | Other specified diabetes mellitus |
| ICD-10 | E13.10 | Other specified diabetes mellitus with ketoacidosis |
| ICD-10 | E13.2 | Other specified diabetes mellitus with renal complications |
| ICD-10 | E13.21 | Other specified diabetes mellitus with established diabetic nephropathy |
| ICD-10 | E13.22 | Other specified diabetes mellitus with end-stage renal disease [ESRD] |
| ICD-10 | E13.3 | Other specified diabetes mellitus with ophthalmic complications |
| ICD-10 | E13.31 | Other specified diabetes mellitus with preproliferative retinopathy |
| ICD-10 | E13.311 | Other specified diabetes mellitus with unspecified diabetic retinopathy with macular edema |
| ICD-10 | E13.319 | Other specified diabetes mellitus with unspecified diabetic retinopathy without macular edema |
| ICD-10 | E13.32 | Other specified diabetes mellitus with proliferative retinopathy |
| ICD-10 | E13.3219 | Other specified diabetes mellitus with mild nonproliferative diabetic retinopathy with macular edema, unspecified eye |
| ICD-10 | E13.3299 | Other specified diabetes mellitus with mild nonproliferative diabetic retinopathy without macular edema, unspecified eye |
| ICD-10 | E13.339 | Other specified diabetes mellitus with moderate nonproliferative diabetic retinopathy without macular edema |
| ICD-10 | E13.3393 | Other specified diabetes mellitus with moderate nonproliferative diabetic retinopathy without macular edema, bilateral |
| ICD-10 | E13.341 | Other specified diabetes mellitus with severe nonproliferative diabetic retinopathy with macular edema |
| ICD-10 | E13.351 | Other specified diabetes mellitus with proliferative diabetic retinopathy with macular edema |
| ICD-10 | E13.3519 | Other specified diabetes mellitus with proliferative diabetic retinopathy with macular edema, unspecified eye |
| ICD-10 | E13.3559 | Other specified diabetes mellitus with stable proliferative diabetic retinopathy, unspecified eye |
| ICD-10 | E13.3599 | Other specified diabetes mellitus with proliferative diabetic retinopathy without macular edema, unspecified eye |
| ICD-10 | E13.39 | Other specified diabetes mellitus with other diabetic ophthalmic complication |
| ICD-10 | E13.40 | Other specified diabetes mellitus with diabetic neuropathy, unspecified |
| ICD-10 | E13.40 | Other specified diabetes mellitus with mononeuropathy |
| ICD-10 | E13.42 | Other specified diabetes mellitus with autonomic neuropathy |
| ICD-10 | E13.42 | Other specified diabetes mellitus with diabetic polyneuropathy |
| ICD-10 | E13.43 | Other specified diabetes mellitus with diabetic autonomic (poly)neuropathy |
| ICD-10 | E13.49 | Other specified diabetes mellitus with other diabetic neurological complication |
| ICD-10 | E13.52 | Other specified diabetes mellitus with certain circulatory complications |
| ICD-10 | E13.621 | Other specified diabetes mellitus with foot ulcer |
| ICD-10 | E13.628 | Other specified diabetes mellitus with other skin complications |
| ICD-10 | E13.65 | Other specified diabetes mellitus with hyperglycemia |
| ICD-10 | E13.69 | Other specified diabetes mellitus with other specified complication |
| ICD-10 | E13.8 | Other specified diabetes mellitus with unspecified complications |
| ICD-10 | E13.9 | Other specified diabetes mellitus without (mention of) complication |
| ICD-10 | E13.9 | Other specified diabetes mellitus without complications |
| ICD-10 | O24.12 | Pre-existing type 2 diabetes mellitus, in childbirth |
| ICD-10 | O24.12 | Pre-existing diabetes mellitus, type 2, in childbirth |
| ICD-10 | O24.33 | Unspecified pre-existing diabetes mellitus in the puerperium |
| SNOMed | 8801005 | Secondary diabetes mellitus |
| SNOMed | 11530004 | Brittle diabetes mellitus |
| SNOMed | 23045005 | Insulin dependent diabetes mellitus type 1A |
| SNOMed | 28032008 | Insulin dependent diabetes mellitus type 1B. |
| SNOMed | 44054006 | Type 2 diabetes mellitus |
| SNOMed | 46635009 | Type 1 diabetes mellitus |
| SNOMed | 73211009 | Diabetes mellitus |
| SNOMed | 81531005 | Type 2 diabetes mellitus in obese |
| SNOMed | 82980005 | Anemia of diabetes |
| SNOMed | 91352004 | Diabetes mellitus due to structurally abnormal insulin |
| SNOMed | 111552007 | Diabetes mellitus without complication |
| SNOMed | 161445009 | H/O: diabetes mellitus |
| SNOMed | 190372001 | Type 1 diabetes mellitus maturity onset |
| SNOMed | 190388001 | Type 2 diabetes mellitus with multiple complications |
| SNOMed | 190389009 | Type 2 diabetes mellitus with ulcer |
| SNOMed | 190447002 | Steroid-induced diabetes |
| SNOMed | 199229001 | Pre-existing type 1 diabetes mellitus.. |
| SNOMed | 199230006 | Pre-existing type 2 diabetes mellitus.. |
| SNOMed | 237599002 | Insulin treated type 2 diabetes mellitus.. |
| SNOMed | 237601000 | Secondary endocrine diabetes mellitus |
| SNOMed | 237604008 | Maturity onset diabetes of the young, type 2.. |
| SNOMed | 237632004 | Hypoglycemic event in diabetes |
| SNOMed | 237633009 | Hypoglycemic state in diabetes |
| SNOMed | 290002008 | Brittle type 1 diabetes mellitus |
| SNOMed | 313435000 | Type 1 diabetes mellitus without complication |
| SNOMed | 313436004 | Type 2 diabetes mellitus without complication |
| SNOMed | 314902007 | Type 2 diabetes mellitus with peripheral angiopathy |
| SNOMed | 314904008 | Type 2 diabetes mellitus with neuropathic arthropathy |
| SNOMed | 315051004 | Diabetes resolved |
| SNOMed | 359642000 | Type 2 diabetes mellitus in nonobese.. |
| SNOMed | 405749004 | Newly diagnosed diabetes |
| SNOMed | 419100001 | Infection of foot associated with diabetes |
| SNOMed | 420270002 | Ketoacidosis in type 1 diabetes mellitus. |
| SNOMed | 420279001 | Renal disorder associated with type 2 diabetes mellitus |
| SNOMed | 420715001 | Persistent microalbuminuria associated with type 2 diabetes mellitus. |
| SNOMed | 420789003 | Diabetic retinopathy associated with type 1 diabetes mellitus |
| SNOMed | 420825003 | Gangrene associated with type 1 diabetes mellitus |
| SNOMed | 421468001 | Neurological disorder associated with type 1 diabetes mellitus |
| SNOMed | 421631007 | Gangrene associated with type 2 diabetes mellitus |
| SNOMed | 421750000 | Ketoacidosis in type 2 diabetes mellitus. |
| SNOMed | 421893009 | Renal disorder associated with type 1 diabetes mellitus |
| SNOMed | 421895002 | Peripheral circulatory disorder associated with diabetes mellitus |
| SNOMed | 421986006 | Persistent proteinuria associated with type 2 diabetes mellitus. |
| SNOMed | 422034002 | Diabetic retinopathy associated with type 2 diabetes mellitus.. |
| SNOMed | 422099009 | Diabetic oculopathy associated with type 2 diabetes mellitus. |
| SNOMed | 426875007 | Latent autoimmune diabetes mellitus in adult |
| SNOMed | 427571000 | Amyotrophy due to type 1 diabetes mellitus |
| SNOMed | 443694000 | Type II diabetes mellitus uncontrolled |
| SNOMed | 444073006 | Type 1 diabetes mellitus uncontrolled |
| SNOMed | 444110003 | Type 2 diabetes mellitus well controlled. |
| SNOMed | 445260006 | Posttransplant diabetes mellitus |
| SNOMed | 472969004 | History of diabetes mellitus type 2 |
| SNOMed | 609561005 | Maturity-onset diabetes of the young.. |
| SNOMed | 609562003 | Maturity onset diabetes of the young, type 1.. |
| SNOMed | 609567009 | Pre-existing type 2 diabetes mellitus in pregnancy.. |
| SNOMed | 703136005 | Diabetes mellitus in remission |
| SNOMed | 713457002 | Neovascular glaucoma due to diabetes mellitus |
| SNOMed | 713702000 | Gastroparesis due to type 1 diabetes mellitus |
| SNOMed | 713703005 | Gastroparesis due to type 2 diabetes mellitus |
| SNOMed | 713704004 | Gastroparesis due to diabetes mellitus |
| SNOMed | 713706002 | Polyneuropathy due to type 2 diabetes mellitus |
| SNOMed | 3321322011 | Type 2 diabetes mellitus uncontrolled |
| SNOMed | 3332232010 | Diabetic ulcer of left foot due to diabetes mellitus type 2 |
| SNOMed | 3332263014 | Diabetic ulcer of right foot due to diabetes mellitus type 2 |
| SNOMed | 3332265019 | Diabetes type 2 with diabetic ulcer of right foot |
| SNOMed | 3513595019 | History of diabetes related lower limb amputation |
| SNOMed | 3688488015 | Kidney disorder due to diabetes mellitus |
| SNOMed | 3688501014 | Peripheral vascular disorder due to diabetes mellitus |
| SNOMed | 3688520011 | Cataract due to diabetes mellitus |
| SNOMed | 3688792011 | Retinopathy due to diabetes mellitus |
| SNOMed | 3690183016 | Clinically significant macular edema of left eye due to diabetes mellitus |
| SNOMed | 3690269011 | Preproliferative retinopathy of right eye due to diabetes mellitus |
| SNOMed | 3690538013 | Macular edema due to type 1 diabetes mellitus |
| SNOMed | 3690545013 | Macular edema due to type 2 diabetes mellitus |
| SNOMed | 3690600017 | Macular edema due to diabetes mellitus |
| SNOMed | 3695522017 | Peripheral neuropathy with type 2 diabetes |
| SNOMed | 3695673019 | Polyneuropathy due to diabetes mellitus |
| SNOMed | 3695736013 | Mixed sensorimotor polyneuropathy co-occurrent and due to diabetes mellitus |
| SNOMed | 3698400010 | Erectile dysfunction with diabetes mellitus |
| SNOMed | 3698404018 | Gastroparesis with diabetes mellitus |
| SNOMed | 3698425018 | Diffuse exudative maculopathy with diabetes mellitus |
| SNOMed | 3698434011 | Advanced retinal disease with diabetes mellitus |
| SNOMed | 3698440016 | Disorder of eye with type 2 diabetes mellitus |
| SNOMed | 3698455013 | Diabetes type II with amyotrophy |
| SNOMed | 3699410014 | Retinopathy with type 2 diabetes mellitus |
| SNOMed | 3699414017 | Proliferative retinopathy with diabetes mellitus |
| SNOMed | 3700525015 | Vitreous hemorrhage with type 1 diabetes mellitus |
| SNOMed | 3702534017 | Proliferative retinopathy of right eye with diabetes mellitus |
| SNOMed | 3775761019 | Proteinuric nephropathy due to diabetes mellitus |
| SNOMed | 3775767015 | Glomerulopathy due to diabetes mellitus |
| SNOMed | 3775798018 | Dyslipidemia due to type 2 diabetes mellitus |
| SNOMed | 3775800013 | Infection of foot due to diabetes mellitus |
| SNOMed | 3776167019 | Disorder of eye due to diabetes mellitus |
| SNOMed | 3776683015 | Vitreous hemorrhage due to diabetes mellitus |
| SNOMed | 3776688012 | Vitreous hemorrhage of left eye due to diabetes mellitus |
| SNOMed | 3776728016 | Macular edema of right eye due to diabetes mellitus |
| SNOMed | 3776730019 | Macular edema of left eye due to diabetes mellitus |
| SNOMed | 3777787015 | Advanced retinal disease due to diabetes mellitus |
| SNOMed | 3778077010 | Disorder of macula due to diabetes mellitus |
| SNOMed | 3778097017 | Diffuse exudative maculopathy due to diabetes mellitus |
| SNOMed | 3778103012 | Disorder of right macula due to diabetes mellitus |
| SNOMed | 3778970010 | Microaneurysm of retinal artery due to diabetes mellitus |
| SNOMed | 3779262016 | Traction detachment of retina due to diabetes mellitus |
| SNOMed | 3779410016 | Retinopathy due to type 2 diabetes mellitus |
| SNOMed | 3779417018 | Disorder of eye due to type 2 diabetes mellitus |
| SNOMed | 3779914017 | Mild nonproliferative retinopathy due to diabetes mellitus |
| SNOMed | 3779995017 | Mild nonproliferative retinopathy of right eye due to diabetes mellitus |
| SNOMed | 3780068012 | Moderate nonproliferative retinopathy due to diabetes mellitus |
| SNOMed | 3780111013 | Severe nonproliferative retinopathy due to diabetes mellitus |
| SNOMed | 3780158017 | Nonproliferative retinopathy due to diabetes mellitus |
| SNOMed | 3780170018 | Preproliferative retinopathy due to diabetes mellitus |
| SNOMed | 3780380014 | Proliferative retinopathy due to diabetes mellitus |
| SNOMed | 3781802017 | Proliferative retinopathy with neovascularization elsewhere than the optic disc due to diabetes mellitus |
| SNOMed | 3782027011 | Disorder of kidney due to diabetes mellitus |
| SNOMed | 3788922010 | Disorder of nervous system due to diabetes mellitus |
| SNOMed | 3788972012 | Autonomic neuropathy due to diabetes mellitus |
| SNOMed | 3788985010 | Neuropathy due to diabetes mellitus |
| SNOMed | 3789017016 | Disorder of nervous system due to type 2 diabetes mellitus |
| SNOMed | 3789545016 | Chronic painful neuropathy due to diabetes mellitus |
| SNOMed | 3789546015 | Chronic painful polyneuropathy due to diabetes mellitus |
| SNOMed | 3789608017 | Neuropathy due to type 1 diabetes mellitus |
| SNOMed | 3789654014 | Peripheral neuropathy due to type 2 diabetes mellitus |
| SNOMed | 3789743013 | Ulcer of heel due to diabetes mellitus |
| SNOMed | 3789783017 | Neuropathic ulcer of heel due to type 2 diabetes mellitus |
| SNOMed | 3793340016 | Erectile dysfunction due to type 2 diabetes mellitus |
| SNOMed | 3793421011 | Dermopathy due to type 2 diabetes mellitus |
| SNOMed | 3800364017 | Gangrene due to diabetes mellitus |
| SNOMed | 3800386016 | Wet gangrene of foot due to diabetes mellitus |
| SNOMed | 3850123012 | Skin ulcer due to diabetes mellitus |
| SNOMed | 3850595016 | Nonproliferative retinopathy of left eye due to diabetes mellitus |
| SNOMed | 3850596015 | Nonproliferative retinopathy of right eye due to diabetes mellitus |
| SNOMed | 3866399017 | Neuropathic arthropathy due to type 2 diabetes mellitus |
| SNOMed | 3866632018 | Hyperglycemia due to diabetes mellitus |
| SNOMed | 3873415016 | Hypoglycemia due to diabetes mellitus |
| SNOMed | 3873807012 | Pancreatogenic type 3c diabetes mellitus |
| SNOMed | 3947162010 | Ulcer of right foot due to diabetes mellitus |
| SNOMed | 3947164011 | Ulcer of left foot due to diabetes mellitus |
| SNOMed | 3947165012 | Chronic ulcer of right foot due to diabetes mellitus |
| SNOMed | 3950862016 | At risk of ulcer of foot due to diabetes mellitus |
| SNOMed | 4009585011 | Bilateral moderate nonproliferative retinopathy due to diabetes mellitus type 2 |
| SNOMed | 4009966018 | Bilateral mild nonproliferative retinopathy due to diabetes mellitus type 1 |
| SNOMed | 4164752010 | Glaucoma suspect due to diabetes mellitus type 2 |
| SNOMed | 4544259019 | Peripheral autonomic neuropathy due to diabetes mellitus |
| SNOMed | 4571144010 | T2DM - diabetes mellitus type 2 |
| SNOMed | 701000119103 | Mixed hyperlipidemia associated with type 2 diabetes mellitus. |
| SNOMed | 721000119107 | Chronic kidney disease stage 4 due to type 2 diabetes mellitus |
| SNOMed | 731000119105 | Chronic kidney disease stage 3 associated with type 2 diabetes mellitus.. |
| SNOMed | 741000119101 | Chronic kidney disease stage 2 associated with type 2 diabetes mellitus. |
| SNOMed | 751000119104 | Chronic kidney disease stage 1 associated with type 2 diabetes mellitus.. |
| SNOMed | 761000119102 | Diabetic dyslipidemia associated with type 2 diabetes mellitus. |
| SNOMed | 771000119108 | Chronic renal impairment associated with type 2 diabetes mellitus. |
| SNOMed | 1481000119100 | Diabetes mellitus type 2 without retinopathy. |
| SNOMed | 1491000119102 | Diabetic vitreous hemorrhage associated with type 2 diabetes mellitus |
| SNOMed | 1501000119109 | Proliferative diabetic retinopathy associated with type 2 diabetes mellitus |
| SNOMed | 1511000119107 | Diabetic peripheral neuropathy associated with type 2 diabetes mellitus. |
| SNOMed | 1521000119100 | Foot ulcer due to type 2 diabetes mellitus |
| SNOMed | 1551000119108 | Nonproliferative diabetic retinopathy associated with type 2 diabetes mellitus.. |
| SNOMed | 1561000119105 | Diabetic peripheral neuropathy associated with type 1 diabetes mellitus.. |
| SNOMed | 1571000119104 | Mixed hyperlipidemia due to type 1 diabetes mellitus |
| SNOMed | 18521000119106 | Microalbuminuria due to type 1 diabetes mellitus |
| SNOMed | 59801000119102 | History of small vessel disease due to diabetes mellitus |
| SNOMed | 71701000119105 | Hypertension in chronic kidney disease due to type 1 diabetes mellitus |
| SNOMed | 71721000119101 | Nephrotic syndrome due to type 1 diabetes mellitus |
| SNOMed | 71791000119104 | Peripheral neuropathy due to type 1 diabetes mellitus |
| SNOMed | 72041000119103 | Osteomyelitis due to type 1 diabetes mellitus |
| SNOMed | 90731000119103 | Chronic kidney disease stage 2 due to type 1 diabetes mellitus |
| SNOMed | 90741000119107 | Chronic kidney disease stage 3 due to type 1 diabetes mellitus |
| SNOMed | 90751000119109 | Chronic kidney disease stage 4 due to type 1 diabetes mellitus |
| SNOMed | 90761000119106 | Chronic kidney disease stage 5 due to type 1 diabetes mellitus |
| SNOMed | 90781000119102 | Microalbuminuria due to type 2 diabetes mellitus |
| SNOMed | 97331000119101 | Macular edema and retinopathy due to type 2 diabetes mellitus |
| SNOMed | 105401000119101 | Diabetes mellitus due to pancreatic injury |
| SNOMed | 111231000119109 | Dyslipidemia with high density lipoprotein below reference range and triglyceride above reference range due to type 2 diabetes mellitus |
| SNOMed | 137931000119102 | Hyperlipidemia due to type 2 diabetes mellitus |
| SNOMed | 138811000119100 | Complication due to secondary diabetes mellitus |
| SNOMed | 138911000119106 | Mild nonproliferative retinopathy due to type 2 diabetes mellitus |
| SNOMed | 140121000119100 | Hypertension in chronic kidney disease stage 3 due to type 2 diabetes mellitus |
| SNOMed | 157141000119108 | Proteinuria due to type 2 diabetes mellitus |
| SNOMed | 164881000119109 | Foot ulcer due to type 1 diabetes mellitus |
| SNOMed | 164971000119101 | Type 2 diabetes mellitus controlled by diet |
| SNOMed | 367991000119101 | Hyperglycemia due to type 1 diabetes mellitus |
| SNOMed | 368051000119109 | Hyperglycemia due to type 2 diabetes mellitus |
| SNOMed | 368551000119104 | Dyslipidemia due to type 1 diabetes mellitus |
| SNOMed | 368581000119106 | Neuropathy due to type 2 diabetes mellitus |

**Appendix S3. Definitions of Covariates**

**Age:** We defined age at baseline based on the underlying variable BirthYear in the raw data. The difference in years from the start of the pandemic and the birth year produced the age at baseline, which was then grouped as 18-39 years, 40-59 years, 60+ years.

**Sex:** We defined sex as the incarcerated resident’s legal gender as recorded in the raw data at the time of their birth (which is a known limitation for the case of a change of legal gender). This variable does not necessarily correspond to gender identity or morphological sex. Male is coded as 1 and Female as 0.

**Race/Ethnicity:** We defined the race/ethnicity of an incarcerated resident based on the Race variable recorded in the raw data. The raw variable has 10 values (Asian/Pacific Islander (a); Black (b); Cuban (c); D - Pacific Islander (d); Hispanic (h); American Indian/Alaskan Native (i); Mexican (m); Other (o); Unknown (u); White (w)). We map these values to 4 categories:

1. Black (Race = Black)
2. Hispanic (Race in {Hispanic, Cuban, Mexican})
3. White (Race = White)
4. Other/Unknown (Race in {Asian/Pacific Islander, D – Pacific Islander, American Indian/Alaskan Native, Other, Unknown})

**COVID-19 Testing Rate:** We defined the rate of COVID-19 testing prior to a positive COVID-19 as follows. We counted all COVID-19 tests recorded in the raw data with sample collection date prior to the date when the first sample that tested positive was collected and on or after March 1, 2020. For individuals who never tested positive, instead of using the first positive sample date, we used the censor date for this interval. To compute the rate, we divided the number of COVID-19 tests by the number of days in the interval and then multiply by 365 to produce an annual rate.

**Historical Medical Use Rate:** We defined the rate of medical use prior to the start of the pandemic as follows. We counted the total number of medical visits of any type for all days in custody between January 1, 2019 and February 29, 2020 and divided by the number of days in custody in this interval and multiply by 365 to produce an annual rate.

**Baseline BMI:** We wished to categorize body mass index (BMI) of incarcerated individuals as close to the start of the pandemic (March 1, 2020) as possible. To do so, for each incarcerated individual, we obtained maximum height values on all days they are recorded in the raw data. We removed any height values that are outliers (<1.219 meters or >2.134 meters). An individual was then assigned the maximum height value across all his/her/their maximum height values recorded. Then, for each incarcerated individual, we obtained raw maximum weight values (kg) on all days they are recorded in the raw data. We removed any weight values that are outliers (<40.823 kg [90 lbs] or >181.437 kg [400 lbs]). For each day with a weight measurement, we calculated body mass index (BMI) as weight/height^2 (kg/m^2). We then removed outlier BMI values (<16.5 or >50). Finally, for each incarcerated individual, we started from February 29, 2020 and moved backwards day by day (up to 12 months prior) until we identified a day with BMI recorded. We classified this BMI as follows:

1. Non-Overweight/Obese: < 25
2. Overweight: 25 to <30
3. Obesity: 30+

**Baseline Blood Glucose:** We wished to categorize blood glucose of incarcerated individuals as close to the start of the pandemic (March 1, 2020) as possible. For each incarcerated individual, we started from February 29, 2020 and moved backwards day by day (up to 12 months prior) until we identified a day where at least one fasting plasma glucose (FPG) test was recorded. Using this date of the closest FPG test, we then looked back an additional 3 months from this date and identified all FPG tests recorded during this 3-month time window. We then took the average of all FPG tests during this 3-month time window and classified these averages as follows for baseline blood glucose:

1. < 70 mg/dL
2. 70-99 mg/dL
3. 100-125 mg/dL
4. ≥126 mg/dL

**Appendix S4. Procedures to predict cumulative risks of diabetes for the population and subpopulation included in the analysis**

We predicted the probability of survival (no incidence of diabetes) over time for all combinations of covariates (covariate profiles) from the selected Cox proportional-hazards model using the survfit() function from the “survival” package in R. We then inverted and linearly interpolated the resulting survival curves using the approxfun() function from the “stats” package in R. We generated 5,000 random survival probabilities (uniformly distributed from 0 to 1) to simulate survival times using the interpolated function. This was done for each covariate profile’s survival curve as well as the 95% confidence interval lower and upper bounds. This resulted in each covariate profile having 5,000 simulated survival times (follow up days), as well as the 95% confidence interval lower and upper bounds.

We then computed the weights for each covariate profile by taking the proportion of

unique residents with that covariate profile out of all unique residents in our observed data. There were cases where no residents in our observed data had a particular covariate profile, in which case the weight of an unmatched covariate profile was calculated to be zero.

We then stratified the simulated survival dataset by group status (SARS-CoV-2 Infection vs No Infection). For each stratum, we calculated the proportion of the total weighted population still at risk for incident diabetes out of the total weighted initial population to determine estimated survival probability at each time point (in days) from 0 (where survival probability was set to 1) to the maximum observed follow up time in our data. This was also done for the 95% confidence interval lower and upper bounds for each stratum. To get the final cumulative hazard curve, we plotted 1 - the survival probability estimates (along with the lower and upper bounds) against time in days (from 0 to the maximum observed follow up days) for each stratum (**Figure 1**).

If the Cox model with interactions was selected, we repeated this process of predicting cumulative risks of diabetes by further stratifying each group (SARS-CoV-2 Infection vs. No Infection) by baseline BMI status and baseline blood glucose level, as well as age and baseline blood glucose level.

For each cumulative hazard curve we predicted, we also extracted the estimated cumulative risk at 1 year (365 days) and 2 years (750 days), along with the 95% confidence interval lower and upper bounds (**Appendix Tables 2-4**).

**Appendix Figure 2. Flow Diagram of Inclusion in the Case-Control Cohort Time-to-Event Study: COVID-19 Test Positive Date Starts Post-Exposure Observation Time**

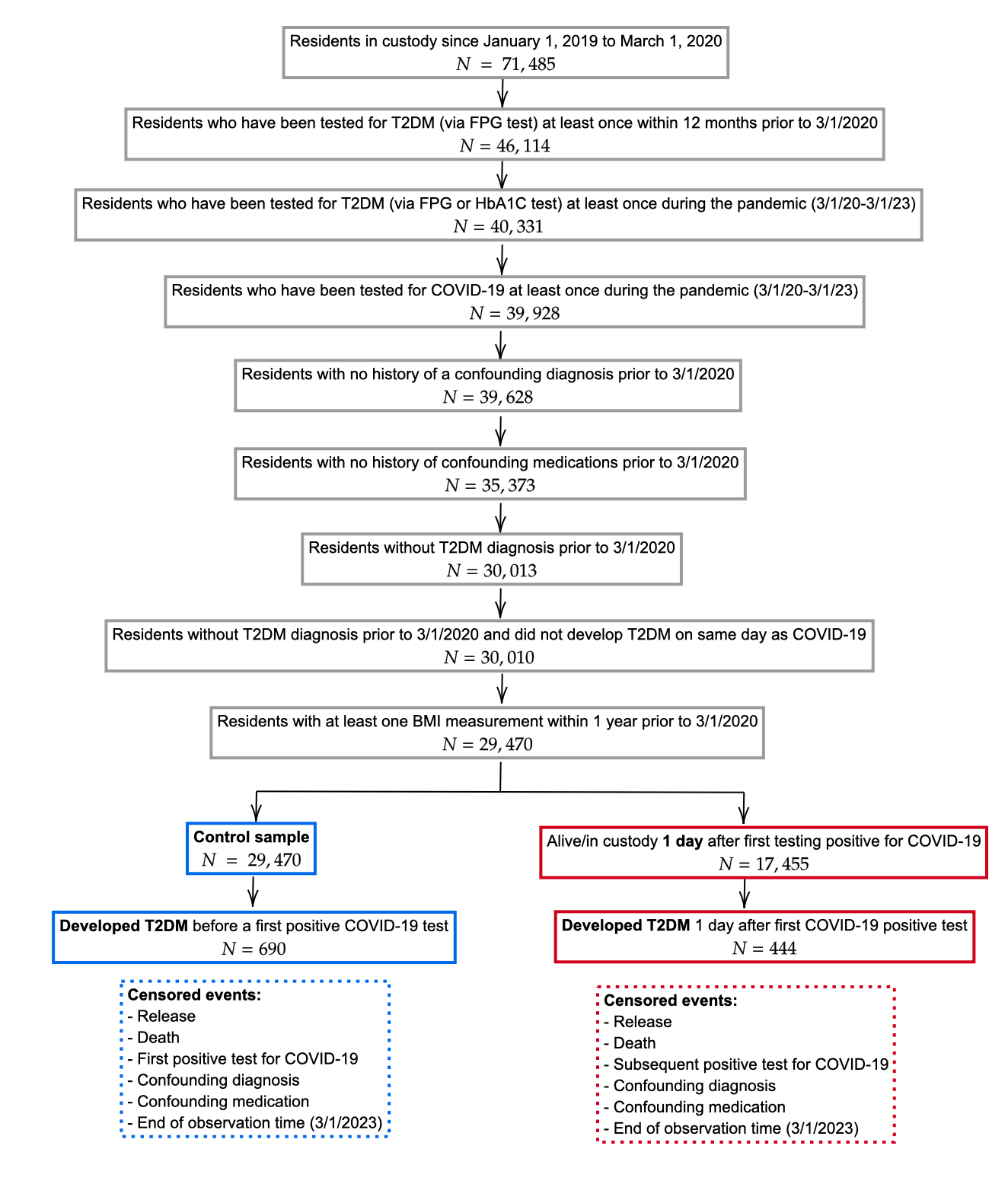

The figure shows the inclusion/exclusion criteria for individual in the study cohort. All cohort members began by contributing (non-exposure) observation time. Individuals who had their first positive COVID-19 test prior to being diagnosed with T2DM (a subset of the control/non-exposure sample) began contributing post-exposure time on the day this test sample was collected (we did not exclude any days from the analysis).

**Appendix Figure 3. Flow Diagram of Inclusion in the Case-Control Cohort Time-to-Event Study: 61 Days after COVID-19 Test Positive Date Starts Post-Exposure Observation Time**

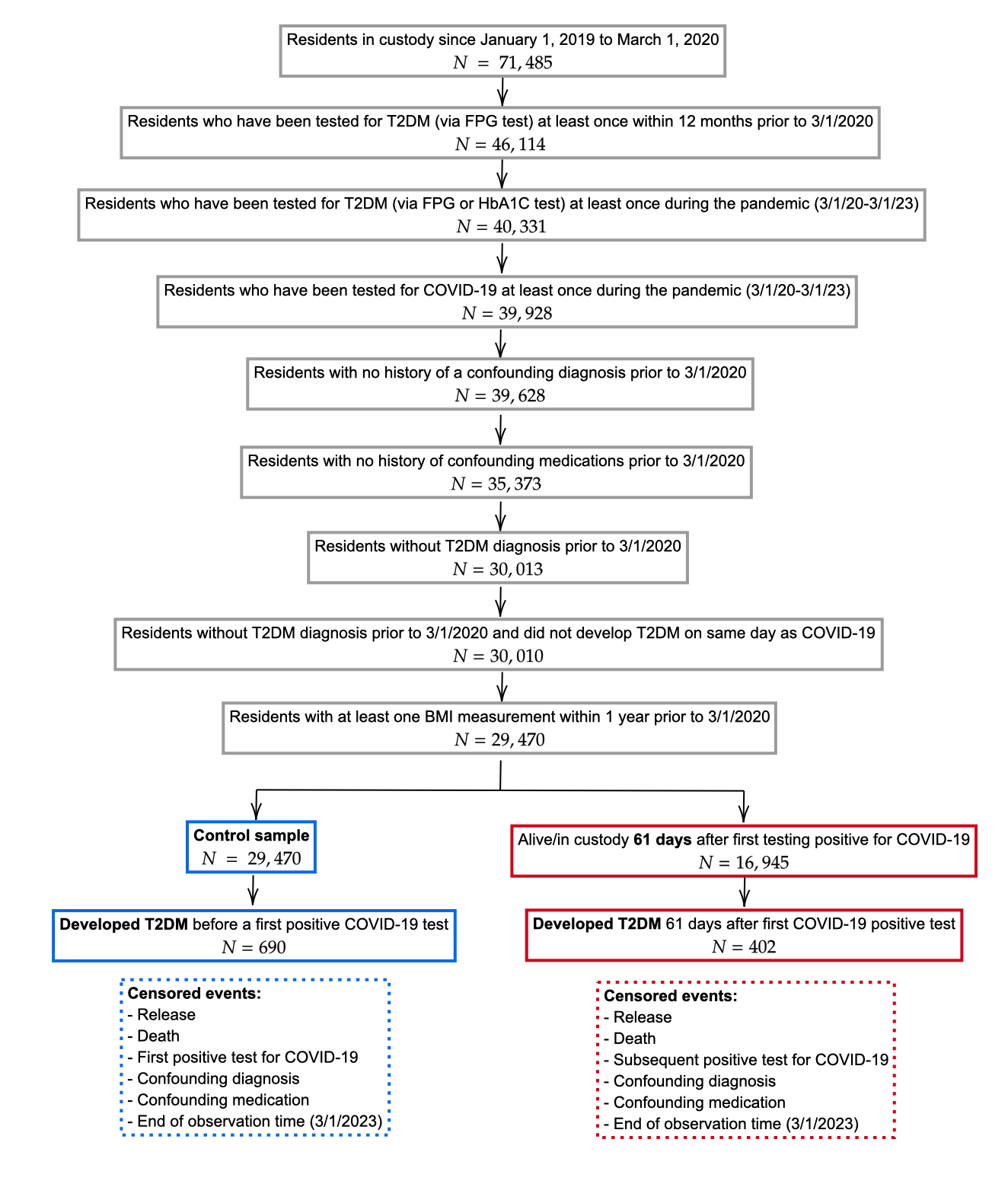

The figure shows the inclusion/exclusion criteria for individual in the study cohort. All cohort members began by contributing (non-exposure) observation time. Individuals who had their first positive COVID-19 test prior to being diagnosed with T2DM (a subset of the control/non-exposure sample) began contributing post-exposure time at 61 days after this test sample was collected (we excluded the first 60 days from the analysis).

**Appendix Figure 4. Flow Diagram of Inclusion in the Case-Control Cohort Time-to-Event Study: 91 Days after COVID-19 Test Positive Date Starts Post-Exposure Observation Time**

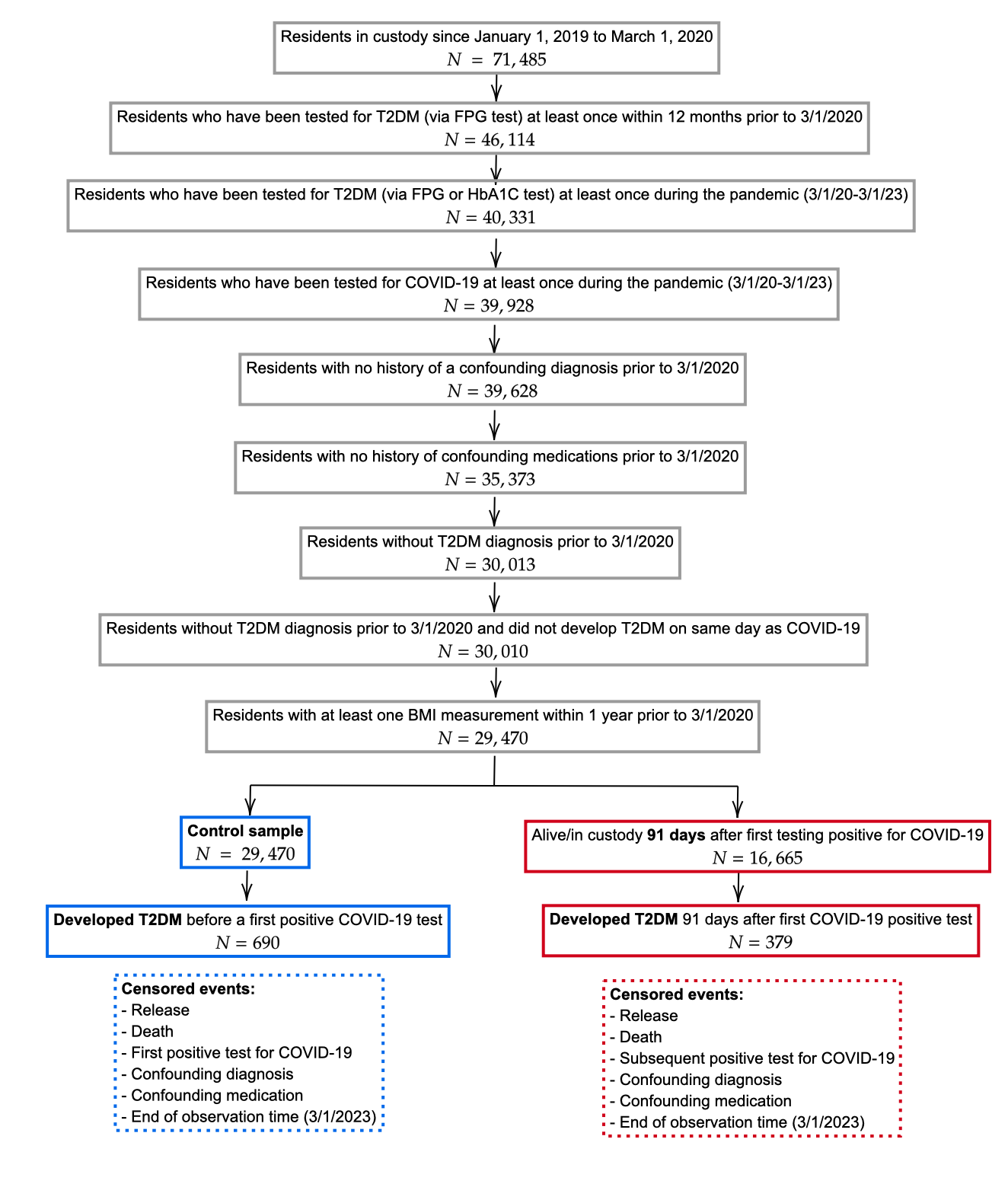

The figure shows the inclusion/exclusion criteria for individual in the study cohort. All cohort members began by contributing (non-exposure) observation time. Individuals who had their first positive COVID-19 test prior to being diagnosed with T2DM (a subset of the control/non-exposure sample) began contributing post-exposure time at 91 days after this test sample was collected (we excluded the first 90 days from the analysis).

**Appendix S5. Assessment of the Potential Confounding Effect of Changes in Diabetes Diagnostic Testing after a Positive COVID-19 Test**

Because the outcome we can observe and analyze is incident *diagnosed* diabetes (Dx DM) and not incident diabetes, it is possible that if COVID infections cause increased rates of DM diagnostic testing, then observed effects of COVID on Dx DM incidence could occur even when COVID has no effect on DM incidence itself. The figure below illustrates this point; Panel A shows a causal model where COVID increases incidence with no effect on diagnostic testing for DM, and Panel B shows a model where COVID has no effect on DM incidence but increases rates of diagnostic testing for DM. We describe the details of our approach for assessing these two possibilities within the context of our analysis below.

*Panel A. An effect of COVID-19 on DM incidence but no effect on DM diagnostic testing*

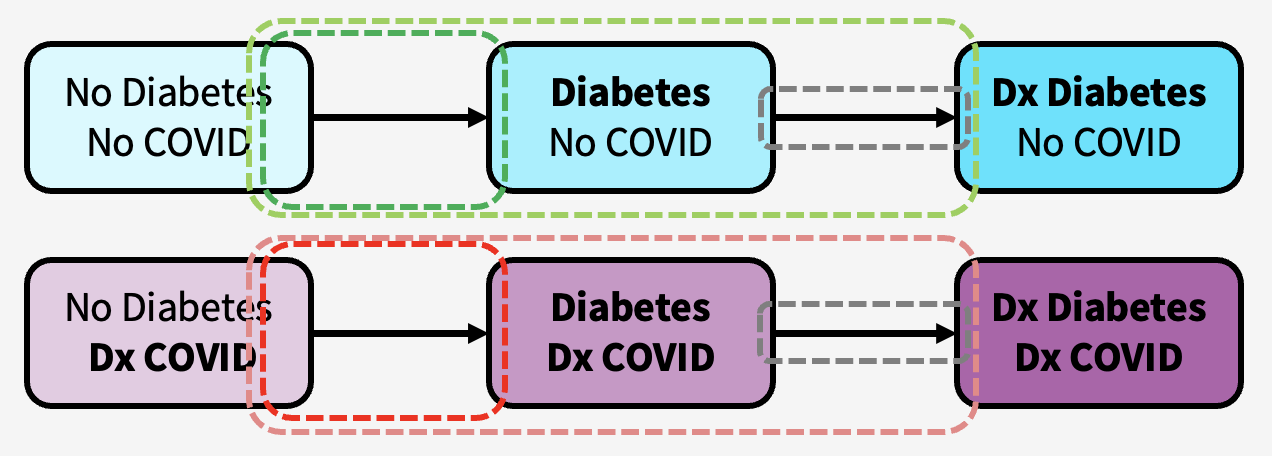

*Panel B. No effect of COVID-19 on DM incidence but an effect on DM diagnostic testing*

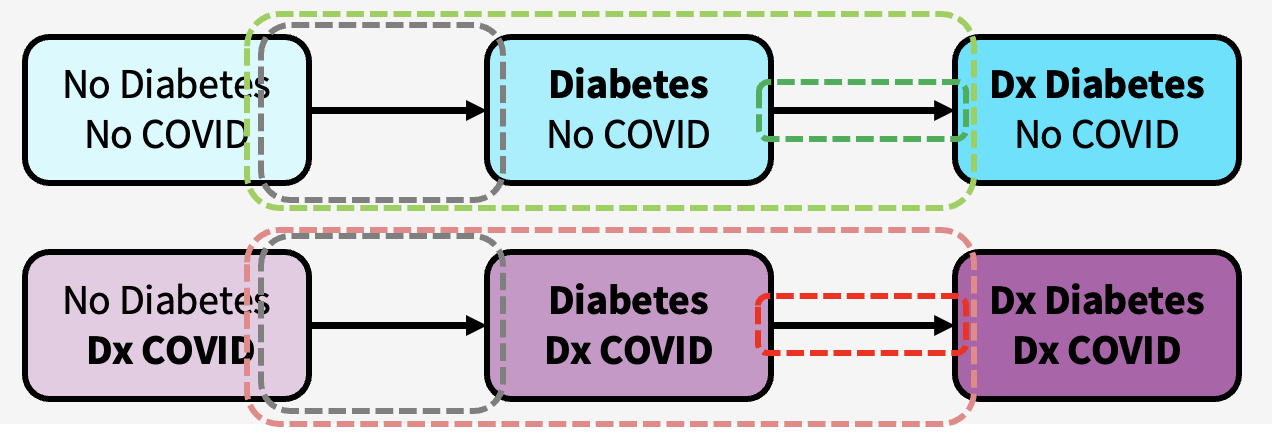

Consistent with the process depicted in figure, we construct a simple simulation model that includes this process for each of two strata (individuals who have not been infected with COVID and individuals after they have been infected with COVID). In each stratum, there are two rates: 1) the rate of developing DM ($r_{1}$); 2) the rate of diagnosis of DM ($r_{2}$). For the stratum for individuals who have been infected by COVID, the model determines these rates with two parameters, thereby relating to the two strata to each other: 1) the hazard rate ratio of DM for people who have been infected with COVID; 2) the hazard rate ratio of diagnosis of DM for people who have been infected with COVID.

For the stratum of people who have not been infected with COVID, the cumulative risk of detected diabetes is

$$F_{NoInf}\left( t \right)=1-\frac{r_{2}e^{-r_{1}t}-r_{1}e^{-r_{2}t}}{\left( r_{2}-r_{1} \right)}$$

For the stratum of people after they have been infected with COVID, the cumulative risk of detected diabetes is

$$F_{COVID19}\left( t \right)=1-\frac{{HRR}_{2}r_{2}e^{-{HRR}_{1}r_{1}t}-{HRR}_{1}r_{1}e^{-{HRR}_{1}r_{2}t}}{\left( {HRR}_{2}r_{2}-{{HRR}_{1}r}_{1} \right)}$$

We apply the simulation model to assess the possibility that confounding due to changes in DM diagnostic testing rates after a COVID-19 infection could explain the differences we estimated in our main analysis. For example, the main analysis produced estimates of the average 2-year cumulative risk of diagnosed diabetes with and without COVID-19 infection (Appendix Table 2): 2.73% without COVID-19 infection and 3.19% with COVID-19 infection.

To do so, we first need to make an estimate of the base rate of DM diagnostic testing among people who had not tested positive for COVID-19 ($r_{2}$) as well as the hazard rate ratio of DM diagnostic testing due to testing positive for COVID-19 (${HRR}_{2}$).

Before differentiating between people with or without COVID-19 infections, we observe that the rate of diagnostic testing for DM (i.e., measuring and recording blood glucose) varied across people in our sample in the pre-pandemic period and during the pandemic. On average people were tested slightly less than every 4 months in both periods. The 25^th^ and 75^th^ percentiles of testing in both periods were approximately every 3.5 months to approximately every 9 months. The rate of DM diagnostic testing declined slightly in the pandemic period.

| Analytic Sample (N=29,470) | 1st Qu. | Median | Mean | 3rd Qu. |
| --- | --- | --- | --- | --- |
| DM test annual rate pre-COVID19 (1/1/2019-2/29/2020) | 1.72 | 2.58 | 2.76 | 3.44 |
| DM test annual rate post-COVID19 (3/1/2020-2/28/2023) | 1.33 | 2.00 | 2.60 | 3.33 |

To estimate the effect of COVID-19 on DM diagnostic testing rates, we use a differences-in-difference approach. For all people who did not test positive for COVID-19, we assign them a “COVID-19 positive” date. We do this by estimating quantile regressions (1,2,…,98,99^th^ quantiles) of the number of days from the start of the pandemic until the date on which the first positive COVID-19 sample was taken, adjusting for all of the covariates used in our main analysis. The quantiles form estimates of the inverse cumulative density function – if we sample uniformly from 0-1, we can determine between which two quantiles we fall for every individual. Then based on their covariate values we can linearly interpolate these quantiles to determine the day on which they were “COVID-19 positive”. The results of this procedure are shown in the figure below.

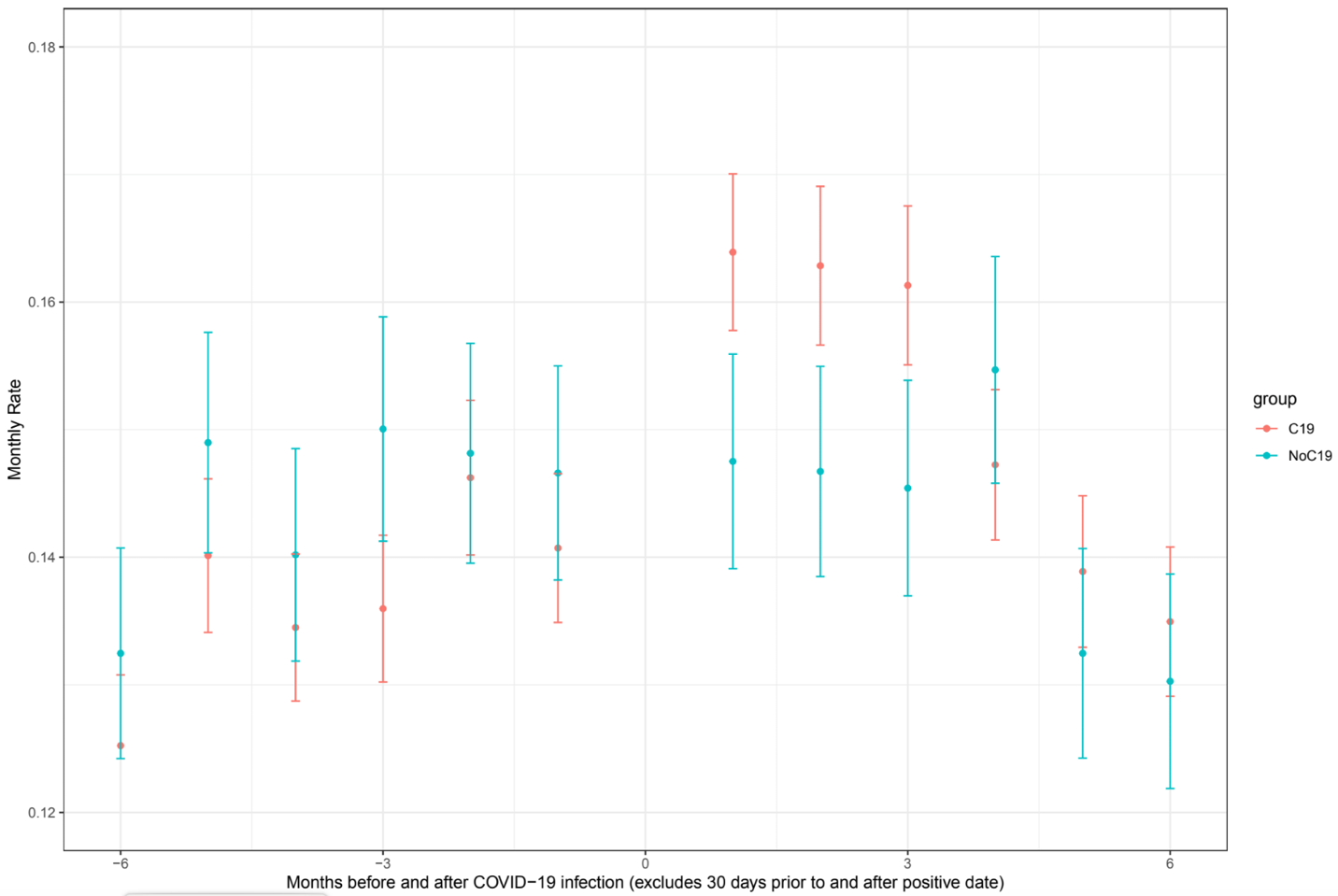

The figure above shows monthly rate 210 to 31 days prior to COVID-19 infection and 31 days to 210 after COVID-19 infection for people who actually had a COVID-19 infection and for those whom we assigned a “COVID-19 positive” date. The crude estimate of the monthly testing rate in the pre-period is 0.146 tests per month in the control group (0.151 in the group of people who had COVID-19 before getting COVID). The crude difference-difference estimate is an increase of about 0.016 tests per month.

|  | Pre | Post | Diff |
| --- | --- | --- | --- |
| COVID-19 | 0.151 | 0.156 | 0.005 |
| Control | 0.146 | 0.167 | 0.021 |
| Diff | -0.06 | 0.011 | 0.016 |

We estimate the difference-in-difference estimate adjusting for baseline covariates and repeat the procedure 1,000 times (i.e., assigning “COVID-19 positive” dates for each person based on random draws based on the quantile regression and then estimating the difference-in-difference regression), producing the results shown in the table below.

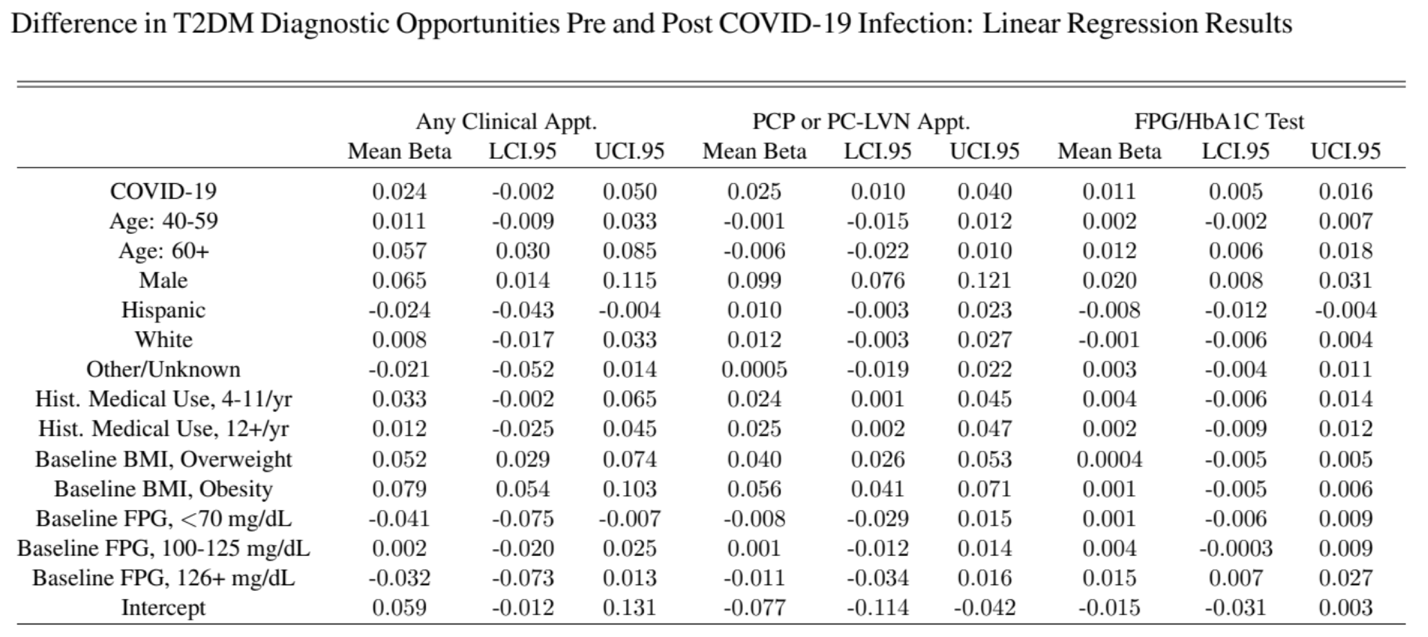

The table above shows results of this procedure for blood glucose testing (FPG/HbA1C test) as well as for clinical appointments overall and for clinical appointments with healthcare providers who would be most likely to measure blood glucose for the purpose of diagnosis. The results show that a COVID-19 infection did not significantly increase the rate of clinical appointments overall or with particular types of providers (after 30 days post-COVID infection – the post-acute period), but COVID-19 did increase DM diagnostic testing rates within these visits by 0.011 tests per month [95%CI: 0.005-0.016]. Given the background testing rates of 0.146 to 0.151 in the pre-period, this implies that ${HRR}_{2}$ could be between 1.03 ([0.151+0.005]/0.151) and 1.11 ([0.146+0.016/0.146).

The above findings along with our main analyses imply that:

1. $r_{2}$ is likely to be 0.004867 DM diagnostic tests per day (0.146 DM diagnostic tests/month)
2. ${HRR}_{2}$ is likely to be about 1.07 and unlikely to be below 1.00 or above 1.21, though we explore values up to 1.50 in the analyses below.
3. ${HRR}_{1}$ is estimated as 1.17 [1.03-1.34] in our main analysis, and we explore values as low as 1.00 (no effect of COVID-19 on HTN incidence) and as high as 1.50 in the analyses below.
4. $r_{1}$(the rate of incident DM among those who have not had COVID-19) is not directly observed but cannot be below the rate of incident diagnosed DM under the assumption that diagnosis is immediate based on the 1-year cumulative rate of diagnosed diabetes in people without COVID-19: 0.0000355/day. A reasonable upper bound might be the rate of incident diagnosed DM among people after COVID-19: 0.0000444. In the analyses below we explore, values as low as 0.000035 and as high as 0.000065.

We fixed $r_{2}$ at its observed value and constructed a dense regular 3-dimensional grid of values in the ranges for $r_{1}$, ${HRR}_{1}$, and ${HRR}_{2}$ listed above (1,370,820 unique combinations). We then used $F_{NoInf}\left( t \right)$and $F_{COVID19}\left( t \right)$to simulate the cumulative risks of incident diagnosed HTN at 1 year and 2 years for each of parameter combinations. We computed sum of squared errors of model-simulated values (cumulative risks of incident detected HTN with and without COVID-19 at 1 year and 2 years and the 1-year and 2-year hazard rate ratios if these cumulative risks were converted to rates under an exponential assumption). We computed the inverse of the sum of squared errors for each parameter combination and normalizing these values such that they summed to 1 across the 1,370,820 combinations. We used these values as weights to sample from these combinations with replacement 2,000,000 times, forming a posterior distribution.

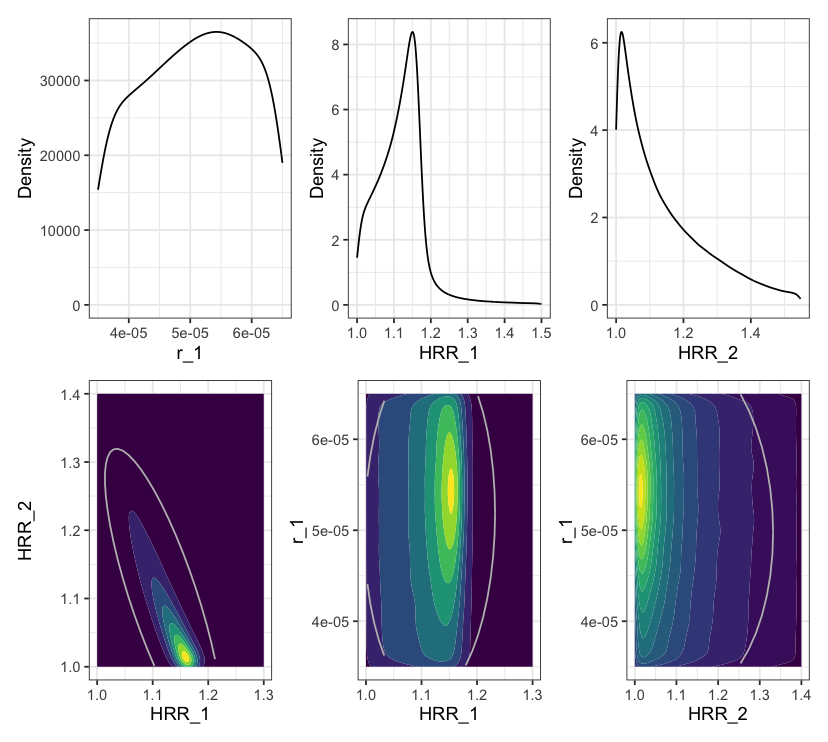

The top row of the figure shows the marginal densities from our procedure – values of $r_{1}$ near 0.000055, values of ${HRR}_{1}$ near 1.15, and values of ${HRR}_{2}$ near 1.025 produce cumulative risks that are most consistent with the observed data. The bottom row shows the relationship between good fitting parameters. In general, if COVID-19 causes a larger increase in the rate of DM incidence (higher values of ${HRR}_{1}$) it must cause smaller increases in the rate of DM diagnostic testing (smaller values of ${HRR}_{2}$). Good fitting values of $r_{1}$ are not strongly correlated with values of either ${HRR}_{1}$ or ${HRR}_{2}$. Again, we can see that it is unlikely that the true effect of COVID-19 on the rate of incident DM is below 1.05 or above 1.20 and that these are consistent with the effect of COVID-19 on the DM diagnostic testing rate of 1.01 to 1.15.

If we plot the 95% and 99% Uncertainty Intervals of ${HRR}_{1}$ conditional on values of ${HRR}_{2}$averaging over the other parameters ($r_{1}$ and $r_{2}$), the following figure results.

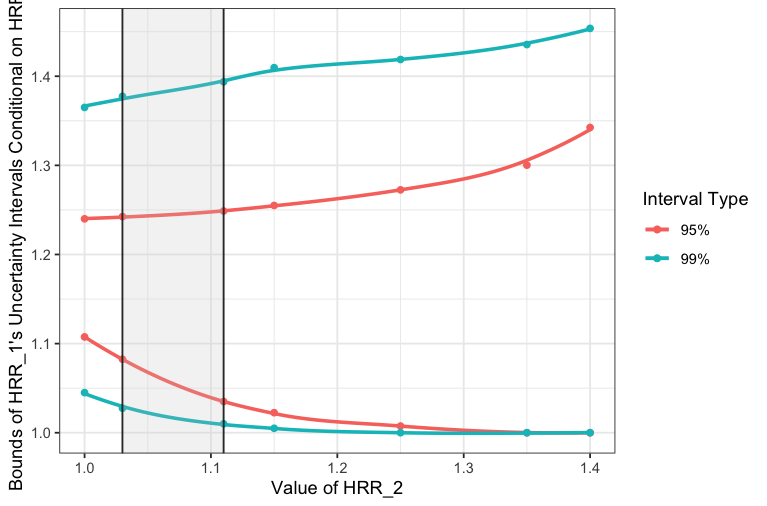

If COVID-19 has no effect on DM incidence, then the lower bounds of the uncertainty intervals (95% or 99%) should include ${HRR}_{1}=1$. This does not occur for values of ${HRR}_{2}$ like those we observed/estimated (generally 1.03 to 1.11, the gray shaded region). In fact, ${HRR}_{2}$ would have to be above 1.35 (1.22) for ${HRR}_{1}$ to have a lower 95% (99%) uncertainty bound that included 1.0 (no effect of COVID-19 on DM).

Finally, we can fix ${HRR}_{2}$ at the estimated value of the effect of COVID-19 on changes in diagnostic testing rate, and then we solve for the smallest value of ${HRR}_{1}$ that makes the model consistent with the differences in the cumulative risk of diagnosed DM at 2 years. If the smallest value of is substantially greater than 1, then we have an estimate of a non-null causal effect of COVID-19 on incident diabetes after removing the confounding effect of changes in diagnostic intensity.

If we fix the underlying rate of incident diabetes for before COVID-19 infection ($r_{1}=0.000054$ per day) and also fix the rate of DM diagnostic testing before COVID-19 infection ($r_{2}=0.000487$ per day) and the increase in DM diagnostic testing post COVID-19 to be ${HRR}_{2}=$1.068, we can then solve for the smallest increase in incident DM due to COVID-19 (${HRR}_{1}$), which is 1.134, substantially greater than a null causal effect. If we repeat this with ${HRR}_{2}=$1.034, ${HRR}_{2}=$1.137, and ${HRR}_{2}=$1.205, our estimates for ${HRR}_{1}$ fall between 1.078 and 1.150.

*Overall Conclusion*

Based on these exercises, we conclude that the confounding effect of increased DM diagnostic testing rates due to COVID-19 may halve the causal effect of COVID-19 on increasing DM incidence (HRR of 1.08-1.10) but is highly unlikely to explain the entire observed effect.

**Appendix Table 1. Results for Estimates and Tests of Significance for Interacted Effects (Estimates are Log HRRs: the Sums of the COVID-19 Main Effect and the Relevant Interaction Effect)**

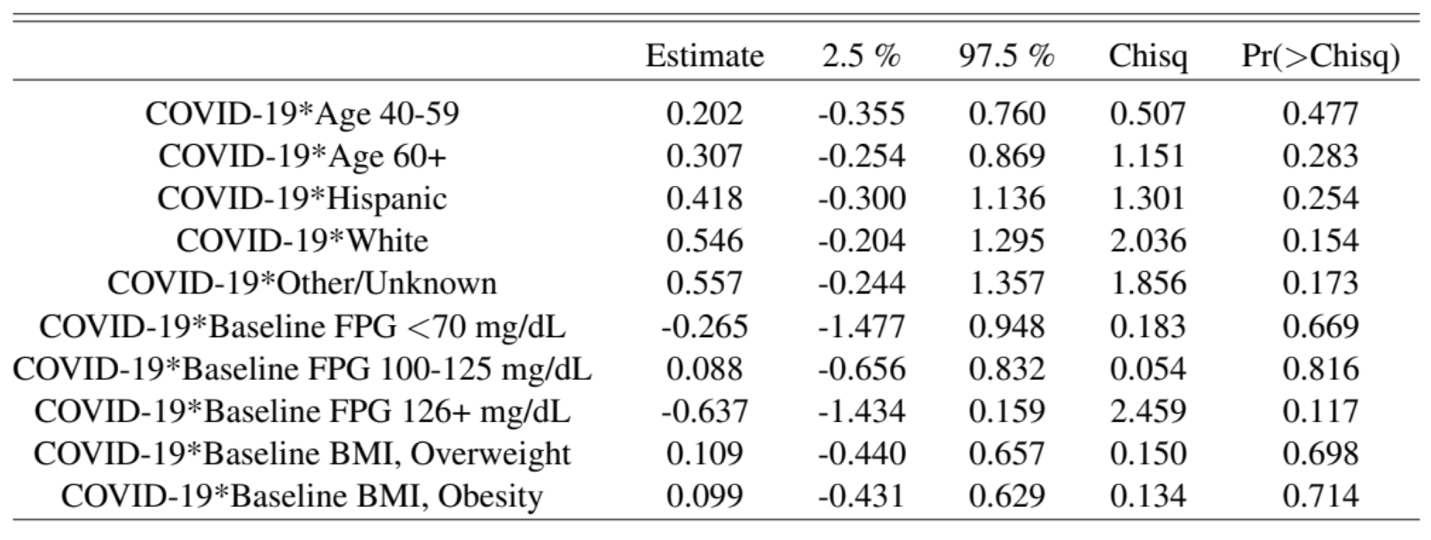

**Appendix Table 2. Predicted Cumulative Risks of Incident Diabetes at 1 Year and 2 Years after Positive COVID-19 Test**

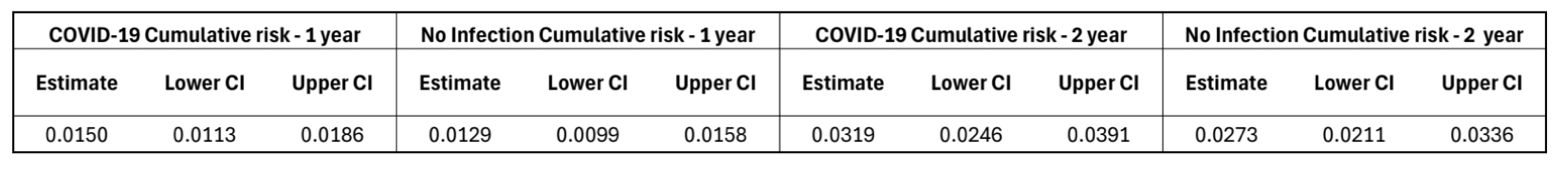

**Appendix Table 3. Baseline Characteristics of Study Population: COVID-19 Test Positive Date Starts Post-Exposure Observation Time**

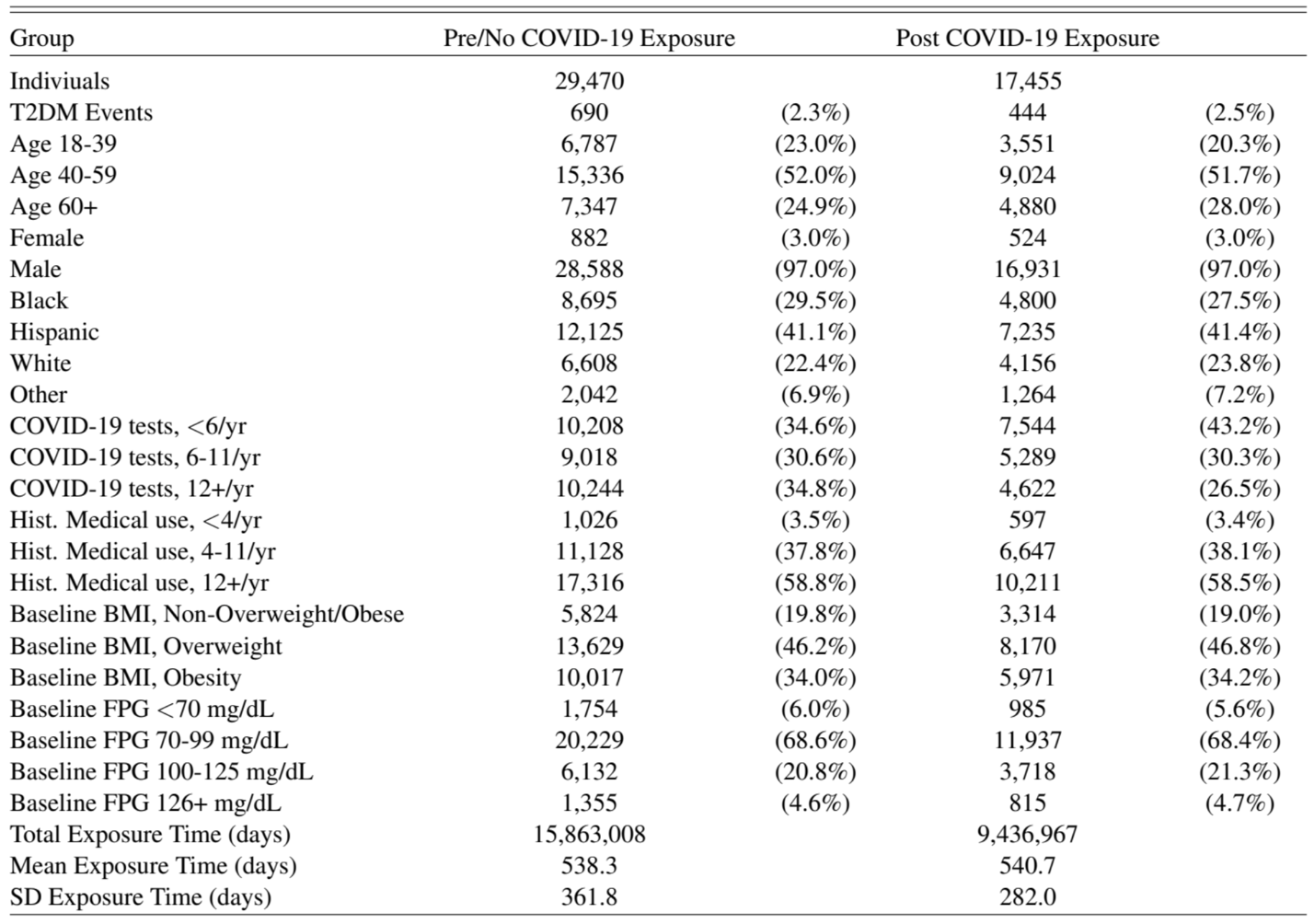

**Appendix Table 4. Baseline Characteristics of Study Population: 61 Days after COVID-19 Test Positive Date Starts Post-Exposure Observation Time**

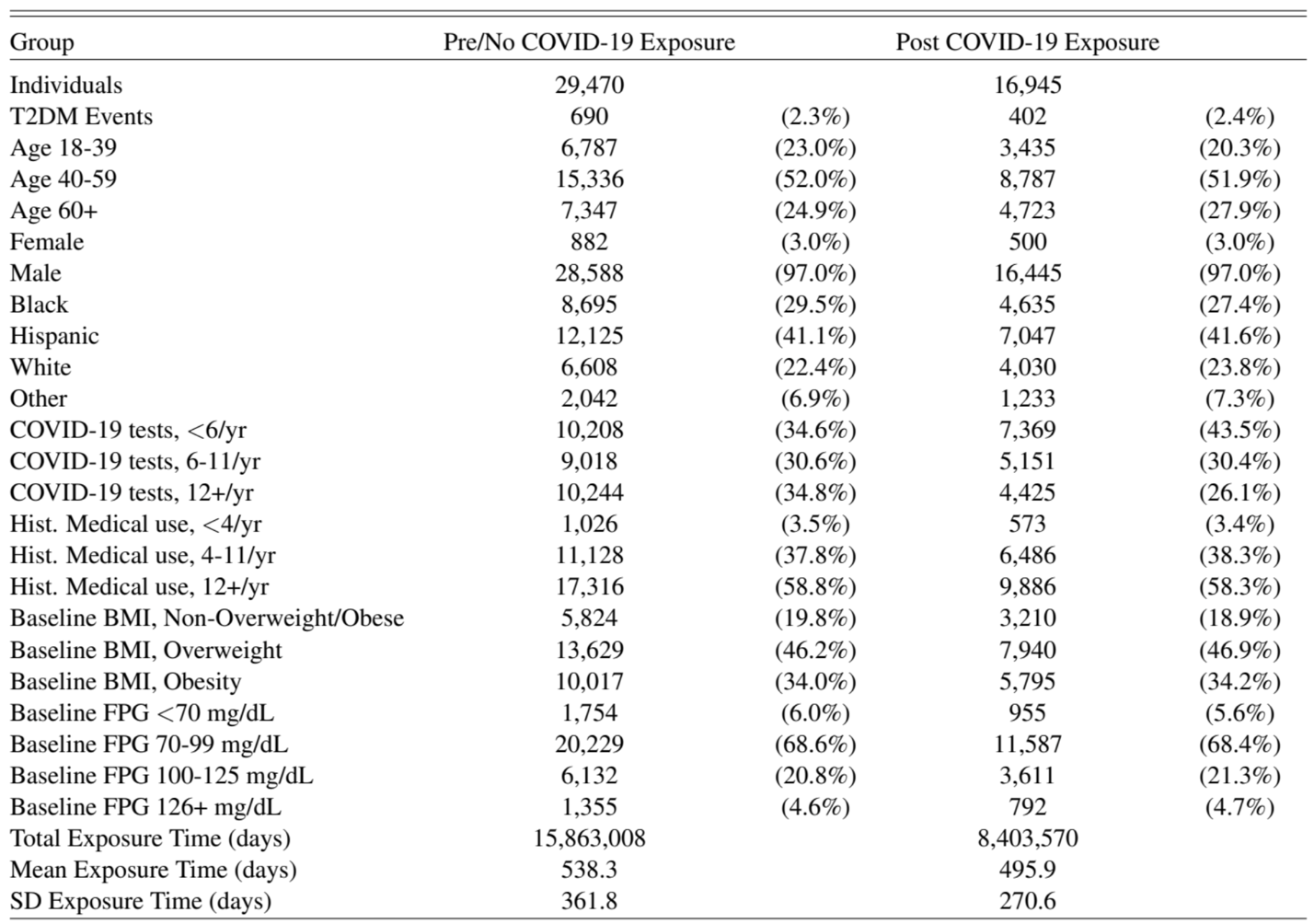

**Appendix Table 5. Baseline Characteristics of Study Population: 91 Days after COVID-19 Test Positive Date Starts Post-Exposure Observation Time**

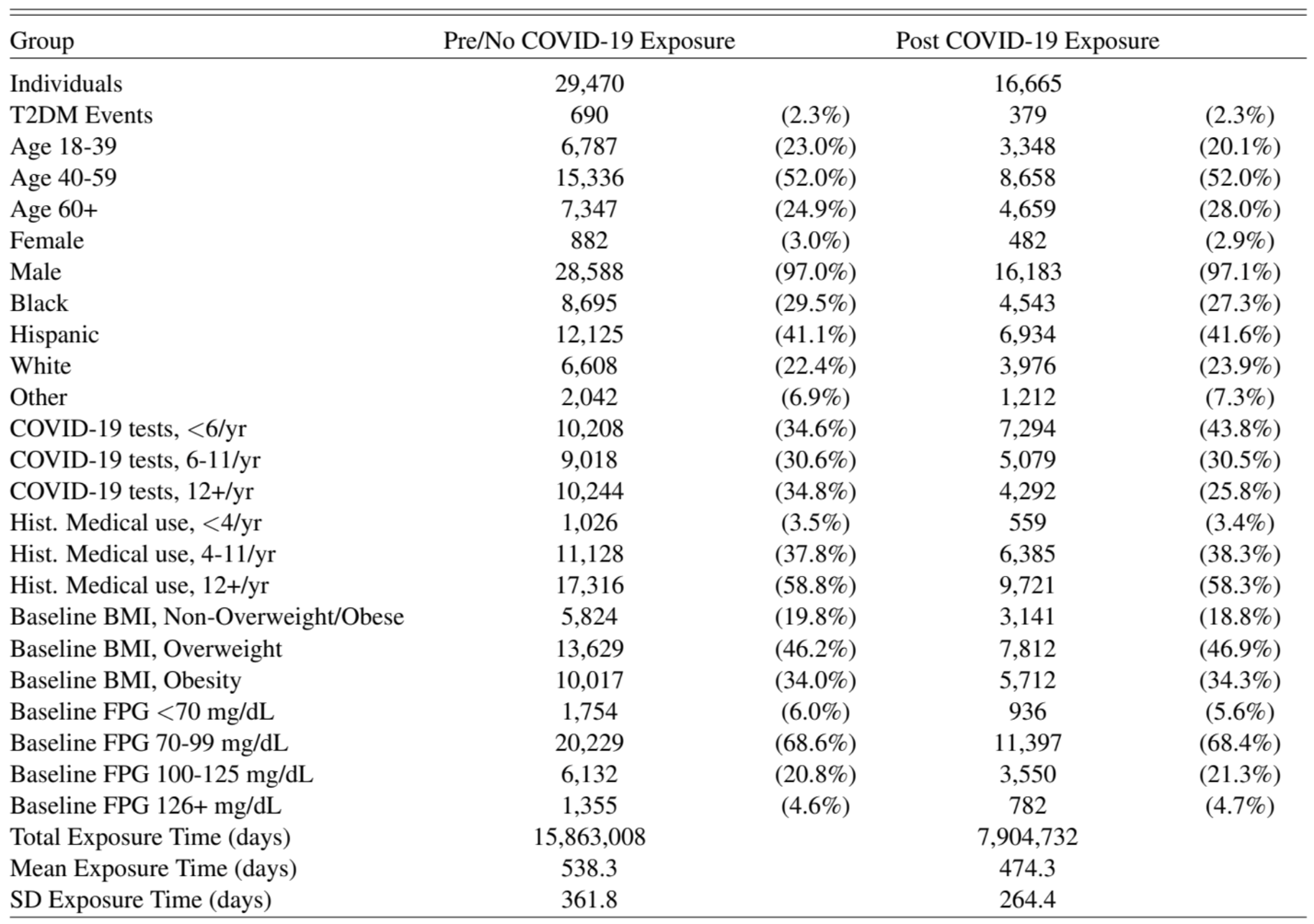

**Appendix Table 6. Multivariate Regression Results for 0-day, 61-day, and 91-day Windows after a Positive COVID-19 Test Starting the Post-exposure Observation Period**

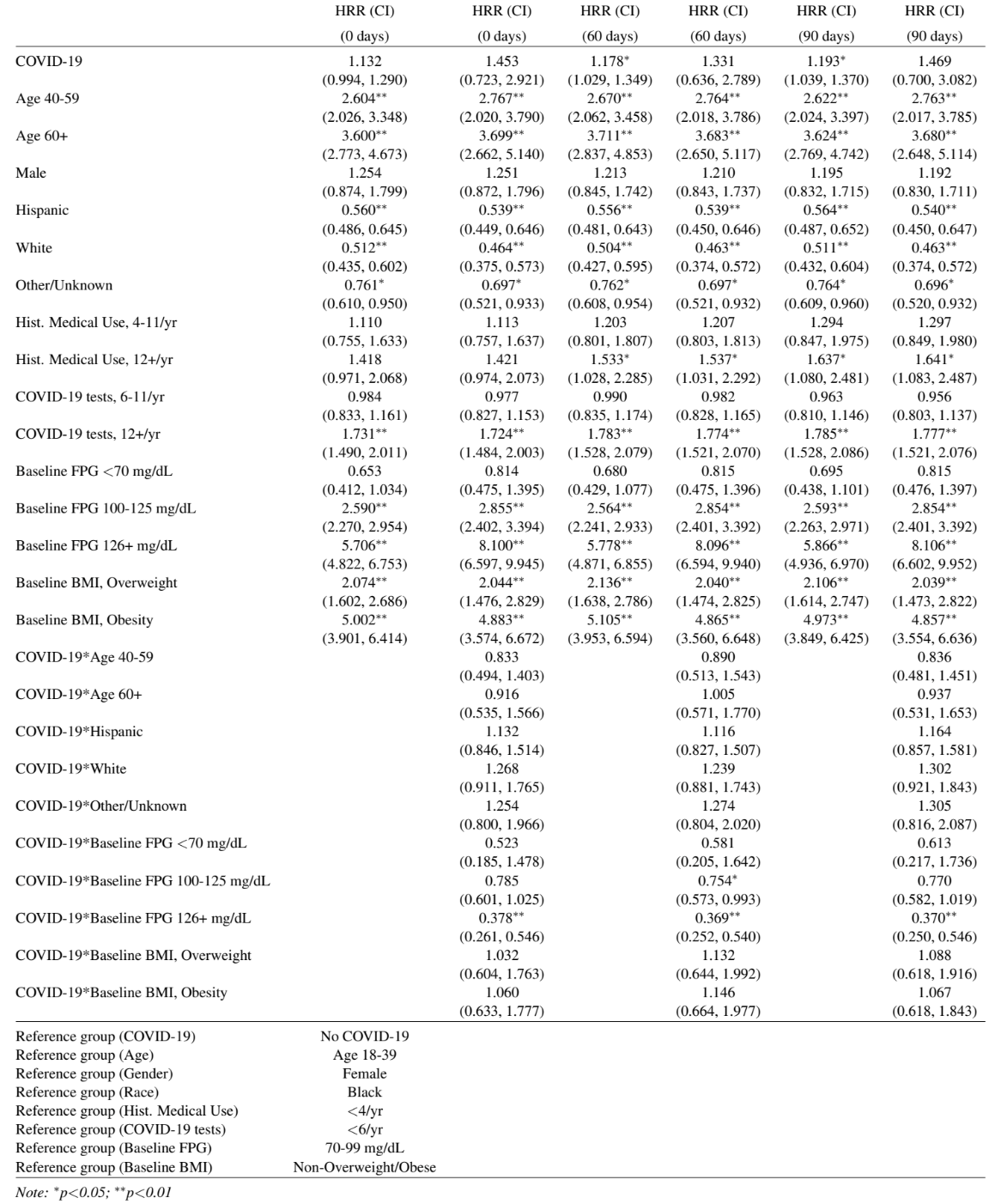
